# Causal modeling of chronic kidney disease in a participatory framework for informing the inclusion of social drivers in health algorithms

**DOI:** 10.1101/2025.11.19.25340498

**Authors:** Agata Foryciarz, Neha Srivathsa, Oshra Sedan, Lisa Goldman Rosas, Sherri Rose

**Affiliations:** Department of Computer Science, Stanford University; Stanford Biobank, Stanford University School of Medicine; Department of Epidemiology and Population Health, Stanford University School of Medicine; Department of Health Policy, Stanford University School of Medicine

**Keywords:** health inequities, social drivers of health, participatory AI, community engaged research, community based systems dynamics, group model building, causal loop diagram, chronic kidney disease

## Abstract

Incomplete or incorrect causal theories are a key source of bias in machine learning (ML) algorithms. Community-engaged methodologies provide an avenue for mitigating this bias through incorporating causal insights from community stakeholders into ML development. In health applications, community-engaged approaches can enable the study of social drivers of health (SDOH), which are known to shape health inequities. However, it remains challenging for SDOH to inform ML algorithms, partially because SDOH variables are known to be interrelated, yet it is difficult to elucidate the causal relationships between them. Community based system dynamics is a community-engaged methodology that can be used to co-create formal causal graphs, called causal loop diagrams, with patients. Here, we used community based system dynamics to create a causal graph representing the impacts of SDOH on the progression of chronic kidney disease, a chronic condition with SDOH-driven health disparities. We conducted focus groups with 42 participants and a day-long model building workshop with 11 participants, resulting in a final graph comprising 16 variables, 42 causal links, and 5 subsystems of semantically related SDOH variables. This final graph, representing the causal relationships between social variables relevant to chronic kidney disease, can inform the development of clinical ML algorithms and other technological interventions.

**Data and Code Availability:** We use data collected from the patient screening survey and questionnaire, as well as notes and recordings generated during focus groups and workshops. The data, beyond summaries reported in the manuscript, are not available to other researchers for privacy reasons. There is no code associated with this work.

**Institutional Review Board (IRB):** This study obtained approval from the Institutional Review Board at Stanford University.

## 1. Introduction

Unfairness and bias in machine learning (ML) algorithms, including in clinical ML algorithms, may be introduced at multiple stages of the algorithm development pipeline (Chen et al., 2021). An underemphasized source of bias is through incomplete or erroneous causal theories, which can lead to biases at the stages of problem formulation or ML algorithm specification (Martin Jr et al., 2020; Prabhakaran and Martin Jr, 2020). Additionally, the causal theories influencing the development of ML algorithms are often implicit, leading to challenges in contesting causal assumptions and identifying sources of bias. Participatory and community-engaged approaches may provide one avenue toward mitigating such bias, through incorporating lived knowledge regarding causal connections from directly impacted community members, such as patients, for clinical ML algorithms.

A number of participatory approaches across fields have engaged patients and their caretakers as partners in health research (Frank et al., 2015; Kimminau et al., 2018; Cohn, 2025), technology development (Bødker et al., 1995; Pilemalm and Timpka, 2008; Donia and Shaw, 2021; Birhane et al., 2022) and policy development (Hovmand, 2014). These approaches include patient centered outcomes research (Frank et al., 2015), community engaged research (Kimminau et al., 2018; Cohn, 2025), communitybased participatory research (Minkler and Wallerstein, 2011), and participatory human-computer interaction and design approaches (Bødker et al., 1995; Pilemalm and Timpka, 2008; Donia and Shaw, 2021; Birhane et al., 2022). Their goal is typically enacting an organizational, policy, or social change for problems pertinent to patient partners. The methods exist along a continuum of engagement, marked by increasing levels of active patient participation, and involve patient partners at different stages of research, from problem identification and scoping, through study or solution design, to evaluation (Birhane et al., 2022; Delgado et al., 2023). They also generate different research outputs, including agreements about changes to the research process, design prototypes, usability tests and knowledge products (such as thematic analysis of user narratives), future visions and needs, and formal conceptual models (Isler and Corbie-Smith, 2012; Dopp et al., 2019; Hansen et al., 2019). Additionally, they have intangible benefits, such as ownership of the research product, increased community capacity, and changed organizational arrangements.

Community-based system dynamics (CBSD) is a community-engaged method that has been advocated for as a path toward mitigating bias in ML algorithms through making causal theories explicit and involving impacted stakeholders at the stage of system understanding and problem formulation (Martin Jr et al., 2020; Prabhakaran and Martin Jr, 2020). For instance, it has previously been used to identify drivers of racial bias in health algorithms (Kuhlberg et al., 2020). CBSD is aimed at modeling non-linear dynamics of complex systems through co-creation with directly impacted community members and originates from the fields of system dynamics and group model building (GMB) (Hovmand, 2014). System dynamics more broadly seeks to model non-linear dynamics of complex systems by generating formal causal representations and quantitative simulations models. GMB is a participatory approach to system dynamics where stakeholder perspectives are combined to generate models. These methodologies have been applied in health settings (Williams et al., 2018; Darabi and Hosseinichimeh, 2020; Smith et al., 2021; Reumers et al., 2022). CBSD utilizes GMB methodology, such as GMB workshops, while emphasizing engagement with community members who have lived knowledge and experience. During a GMB workshop, participants are led through iterative steps of formalizing their understanding of a given system using a common formal graphical vocabulary, leading to the generation of a causal loop diagram (CLD). A CLD can serve as a conceptual representation of a system, helping guide discussion about desired changes and their mechanisms. It can also be further transformed into a simulation model, where dynamics of the system and sensitivity to changes in particular variables can be studied.

CBSD, by formalizing lived knowledge and the experiences of individuals embedded in a given social system, may be especially conducive to eliciting causal theories pertaining to social context. In particular, patients experiencing specific health conditions are uniquely positioned to understand relevant social drivers of persistent health inequities, known as social drivers of health (SDOH). SDOH are “conditions in the environments where people are born, live, learn, work, play, worship, and age that affect a wide range of health … outcomes.” (Pronk et al., 2021; HHS, 2024) They encompass a number of domains, including education, health care access, neighborhood, social and community context, and economic stability (Solar and Irwin, 2010; Pronk et al., 2021; HHS, 2024). SDOH additionally comprise two levels: intermediary and structural SDOH. Intermediary SDOH impact patients at the individual and community-level, while structural SDOH are the upstream factors influencing social stratification (Solar and Irwin, 2010).

Although theories of social epidemiology emphasize the interrelated nature of SDOH, empirical studies of the impact of SDOH on health outcomes and disparities often consider only a subset of the SDOH variables within each domain (Solar and Irwin, 2010). This makes it difficult to formalize statistical models that incorporate multiple SDOH. Patients experiencing specific health conditions may have a more complete understanding of relevant SDOH factors and their interactions than researchers. Participatory research approaches, such as CBSD, may therefore be beneficial for generating a system-level understanding that captures mutual relationships between SDOH variables within and across domains and levels. Such an understanding can enable the design of technology and policy aimed at reducing health inequities attributable to those drivers.

Persistent health disparities are particularly pronounced for chronic conditions, such as chronic kidney disease (CKD). CKD is highly heterogenous in its origins and presentation, and often co-occurs with other chronic conditions like diabetes and cardiovascular disease. Kidney damage caused by CKD is irreversible, making early diagnosis and appropriate management critical. Clinical management of CKD encompasses periodic monitoring, comorbidity management, and preparing for renal replacement therapy in later stages. Non-clinical management includes lifestyle adjustments related to exercise, diet, water intake, and the management of physical symptoms (Levin et al., 2013; Wouters et al., 2015; Shlipak et al., 2021).

In the U.S., patients who are Black, Asian, Hispanic, Native American, and from lower socioeconomic backgrounds have higher rates of CKD disease progression and end stage renal disease compared to white patients (Narva, 2008; Saran et al., 2017; Ahmed et al., 2021). Documented SDOH contributing to these disparities include access to and quality of medical care, presence of social support, and access to healthy food and walkable spaces (Navaneethan et al., 2008; Crews et al., 2014; Norton et al., 2016; Chu et al., 2021; Hannan et al., 2021; Eneanya et al., 2022). Varied methodological approaches have been applied to study factors impacting the health of individuals with CKD and their experience of navigating care and self-management (Crews et al., 2014; Norton et al., 2016; Kang et al., 2017; Teasdale et al., 2017; Kang et al., 2018; Hannan et al., 2021; Eneanya et al., 2022). System dynamics approaches have been applied to study CKD (Motohashi and Nishi, 1991; Kang et al., 2017, 2018) and SDOH (Williams et al., 2018; Smith et al., 2021; Reumers et al., 2022). However, to our knowledge, there has been no prior participatory research involving CKD patients to generate a formal system-level understanding of interactions between different SDOH relevant to CKD.

## 2. Group Model Building

During a GMB workshop, a small group of participants is led through a series of structured activities to create a CLD—a formal graphical causal representation of a complex system (Hovmand, 2014). The study team typically includes 3–5 people who have fixed roles throughout the GMB process, including a facilitator, modeler, process coach, and recorder. The structured activities, described by *scripts*, incrementally convert and combine participants’ perspectives into consensus-based, shared visual representations of increasing complexity. The process starts with identifying relevant variables and individual connections between them, and ends in creating a complete CLD and analyzing its structure (Hovmand et al., 2011). The structure is then used to discuss mechanisms through which desired changes in the diagrammed system can be achieved. The entire GMB process can be conducted over a single day or span multiple days. A CLD, the final product of a GMB process, is a graph consisting of variable nodes connected by causal links. In contrast with other causal diagramming methodologies such as directed acyclic graphs, CLDs allow and emphasize feedback loops, which illustrate complex dynamic relationships within a system.

GMB processes often begin with identifying a *main variable*—the key dynamic variable for a problem of interest. Participants are also introduced to core GMB concepts (Figure 1), through the *reference mode* and a *concept model*. The reference mode is a graph showing different possible trajectories of the main variable over time (Figure 1 A). The *concept model* is a small, simple CLD, used to introduce the concepts and iconography of variables, causal links, and causal link *polarity* (Figure 1 D). Causal links are depicted as arrows between pairs of variables, pointing from cause to effect. Each link is marked by a plus or minus sign, indicating whether variables change in the same direction (*positive polarity*) or opposite directions (*negative polarity*) (Figure 1 B). Variables not directly connected by a causal link can still be causally related, if a causal path through *mediating variables* exists between them (Figure 1 C). The concept model can also serve as a seed structure upon which a CLD is built.

**Figure 1.**
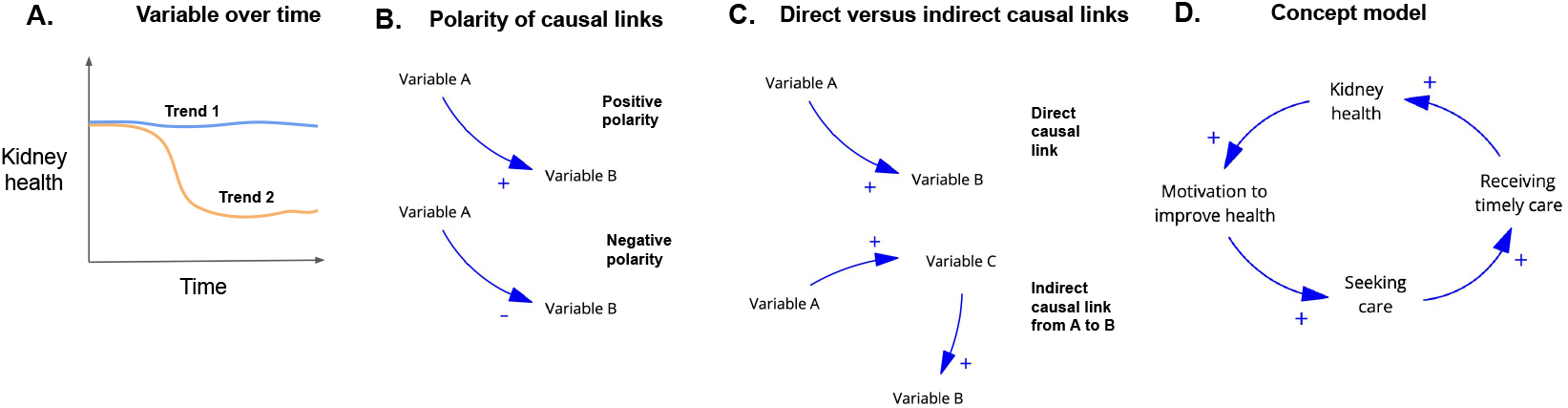
Core group model building concepts introduced to participants during the workshop. (A) Reference mode, showing trajectories of the main variable *kidney health* over time. (B) Polarity of causal links. Positive polarity indicates that variables change in the same direction. Negative polarity indicates that variables change in opposite directions. (C) Direct versus indirect causal links. Variable A may have a direct causal link to variable B, or it may have an indirect causal link to variable B via the mediator variable C. (D) Initial concept model introduced to participants during the first session of the workshop. The link from *kidney health* to *motivation to improve health* was suggested by participants, while the remaining variables and links were introduced by a facilitator.

The next stage involves variable elicitation and refinement, where GMB participants identify variables that influence the main variable. A selected set of variables is used by participants to form connection circles, where pairs of variables are connected by causal links based on participant knowledge and experience. Next, participants construct the CLD, adding variables and causal links to the concept model, using intuition built from forming connection circles. Finally, the structure of the CLD is analyzed and desired system interventions are discussed. Through this process, scripts often alternate between small groups of 3–4 participants and a plenary, where insights are integrated across the multiple small groups. While our study focuses on CLD construction, we note that some implementations of GMB can include participatory quantitative simulation modeling, which follows this qualitative step.

## 3. Methods

### 3.1. Recruitment

The five person study team consisted of faculty, students, and a staff member from the Stanford Schools of Medicine and Engineering. We partnered with the Stanford Participant Engagement Program (PEP) to recruit 50 adult, English-speaking Stanford Health Care patients whose medical records included at least two CKD diagnosis codes since 2017. Stanford Health Care is a large academic health care system in Palo Alto, California, primarily serving patients with employer-based insurance residing in the San Francisco Bay Area. The study team never accessed patients’ medical records and the PEP team identified the cohort of individuals who met inclusion criteria based on information in their medical records. PEP implemented a sampling procedure from this cohort that applied equal weights based on patients’ binary sex, race, and ethnicity information to create a cohort of 2500 potential participants (given the anticipated response rate of about 2%). Potential participants were sent physical letter invitations. Patients indicated interest in the study by completing an online screening survey via a link provided in the letter or calling a number provided in the letter and providing information to a study team researcher who completed the screening survey on their behalf. Patients were recruited to focus groups, and focus group participants were invited back to participate in a full-day workshop. Participants were offered payment for taking part in the study: $100 for participating in a focus group and $330 for participating in the full-day workshop. Enrolled patients filled out an additional survey that asked about their race and ethnicity, CKD stage, education level, employment status, marital status, and insurance type. The survey is available in Supplement 2.

### 3.2. Focus Groups

We conducted four 60- to 90-minute focus groups— two in person and two online. The aim of the focus groups was to gain a better understanding of major concerns and themes related to CKD care among the study participants, inform workshop design, and build trust between study participants and the study team ahead of the workshop. The focus group facilitators asked participants questions pertaining to their knowledge about CKD, social and community context, economic stability, and how these factors impacted their CKD, among other topics. A comprehensive list of all focus group questions, from which a subset of questions was used for each focus group, is provided in Supplement 1. Members of the study team aggregated main themes discussed based on notes taken during the focus groups.

### 3.3. Workshop Sessions

Our GMB workshop consisted of 13 activities, each 15- to 30-minutes long, divided amongst four 60- to 90-minute sessions across a single day. The single day format was designed to promote participant retention. The activities were based on scripts adapted from Scriptapedia, with modifications described in the next subsection (Hovmand et al., 2011). The workshop scripts are summarized in Appendix A, and the workshop schedule and scripts are provided in Supplement 4 and Supplement 5, respectively. During the workshops, four team members comprised the study team, with AF and NS as facilitators, NS as the modeler, LGR as the process coach, and SR as the recorder (Richardson and Andersen, 1995). We provide the dictionary mapping technical GMB terminology to participant-facing terminology in Supplement 3.

### 3.4. Development of Scripts

When developing scripts, we sought to account for findings from our focus groups, mock workshop sessions that we conducted with Stanford researchers, and the study goals. We aimed to encourage discussion of a diverse set of variables, spanning a number of SDOH domains and ranging in level from intermediary to structural. To achieve this, we explicitly introduced participants to domains of SDOH at the beginning of the workshop. Additionally, during variable refinement, we clustered variables proposed by participants by SDOH domain and level, to visualize which categories were receiving less attention. We initially excluded the kidney health variable in the small group connection circle script, adding it later in the script. This was to encourage the identification of causal connections between any pair of variables and indirect links between variables and kidney health. There were also script changes implemented on the day of the workshop, due to time constraints. During graph construction, the facilitators introduced a “rapid fire link round” to efficiently debate a series of links and reach consensus about each within a limited time. Another change involved abbreviating three of the last four sessions, including loop and path analysis and the dot exercise for interventions.

### 3.5. Post-Workshop Refinement

Following the workshop, the study team iterated on the CLD generated during the GMB sessions. The changes reflected relationships discussed during focus groups and workshops, by specifying variables and links, and simplifying the resulting diagram. Updates included adding, renaming, and merging variables, as well as adding, removing, and changing the polarity of links. Further analysis of the CLD also involved identifying subsystems within the CLD. The subsystems consisted of semantically related variables with causal links between them that served a functional purpose within the larger CLD.

## 4. Results

### 4.1. Study Sample

Of the 2494 patients invited to the study, 119 contacted us and 72 met inclusion criteria and consented to participate. There were 6 damaged labels and 37 letters returned to us without reaching patients. We enrolled 52 patients into one of four focus groups, 42 of whom participated. Finally, 11 focus group participants took part in the GMB workshop. The flowchart illustrating recruitment is included in Appendix D.

We report demographic summaries of all 42 participants for privacy reasons. Our recruitment process invited a comparable number of men and women to the study, but all participants who enrolled were women. Participants represented stages 2–5 of CKD, and included individuals on dialysis and individuals who received a kidney transplant. Some were not aware of their CKD diagnosis when they received the study invitation, and others did not know their CKD stage. The majority of the women were either Black or African American (n=12) or Asian (n=19), and were 56 years old on average. Close to half were privately insured (n=22), with the rest receiving health insurance through Medicare or MediCal (n=20). Over half of the participants (n=22) had advanced degrees, with 14 holding graduate or professional degrees.

### 4.2. Focus Groups

Main themes that emerged from the focus groups included nutrition and diet, trust and communication with physicians, comorbidities and complex health management, transportation, time and logistics of care, knowledge gaps and CKD awareness, emotional and mental health, social support, inequities in access to resources, and social stigma. A detailed summary of the themes is available in Appendix B.

While awareness of CKD differed across the group, overall we noted a high degree of health literacy, with many patients discussing laboratory tests and their changing results with fluency. This was particularly true among people who had been diagnosed for a long time and whose primary health concern was CKD.

### 4.3. Causal Loop Diagram

The final CLD incorporating updates by the study team is shown in Figure 2. The original CLD generated at the end of the GMB workshop is included in Appendix C. A complete list of modifications made to the original CLD and the rationale for these modifications is available in Supplement 6. The final diagram was composed of 16 variables and 42 links. We identified 5 primary subsystems: care management, health knowledge and information, socioeconomic resources, social support and responsibilities, and health behaviors. The care management and health behaviors subsystems directly impacted *kidney health*, and mediated the impact of the remaining subsystems on *kidney health*.

**Figure 2.**
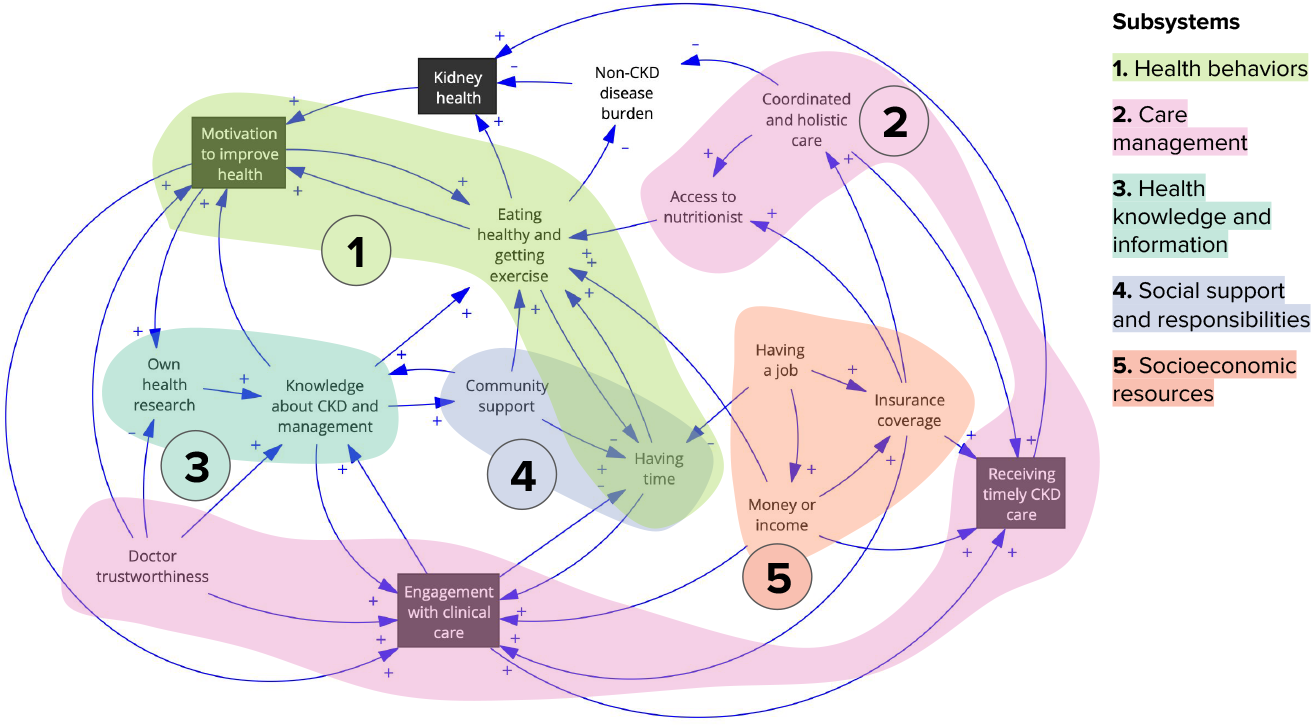
Final causal loop diagram (CLD), with subsystems marked. The CLD consisted of variables connected by causal links, depicted as arrows pointing from cause to effect, with signs indicating positive and negative polarity. The variables originating from the concept model, introduced to participants at the beginning of the workshop, were marked in black boxes. The shaded areas within the CLD marked subsystems, which consisted of semantically related variables that served a functional purpose within the larger CLD.

The health behaviors subsystem consisted of *motivation to improve health, eating healthy and getting exercise*, and *having time*. This subsystem captured the reinforcing relationship between motivation and health behaviors, which are time consuming. It impacted both *kidney health* and *non-CKD disease burden* directly. Participants described the effort nec-essary for making lifestyle adjustments, and challenges with explaining dietary restrictions to friends and family. They also mentioned the importance of knowledge regarding dietary restrictions and fluid intake.

The care management subsystem was composed of *doctor trustworthiness, engagement with clinical care, receiving timely CKD care, coordinated and holistic care*, and *access to nutritionist*. It mediated the impact of the other four subsystems on *kidney health*, showing the range of variables that impacted one’s ability to engage in clinical care. Participants discussed experiences related to *coordinated and holistic care*, including the need for specialists to consider multiple co-occurring conditions and communicate between care team members. They also mentioned the limited nutritional counseling they received from physicians, identifying *access to a nutritionist* as an unmet need among early and moderate-stage CKD patients. Patients in later stages were connected to a nutritionist after reaching stage 5, noting the difference having a nutritionist made in their management journey. The importance of *doctor trustworthiness* was discussed at length. The CLD reflected how *doctor trustworthiness* increased patient *engagement with clinical care*, which improved *knowledge about CKD and management*, while a lack of *doctor trustworthiness* can lead to reliance on their *own health research* for *knowledge about CKD and management. Doctor trustworthiness* also increased *motivation to improve health*. Needs related to *doctor trustworthiness* included ensuring that doctors treat patients with respect, share information pertinent to care, including all relevant diagnoses and management, and do not lead with biases that would lead to a dismissal of symptoms, such as anti-fat bias, medical racism, or ageism.

The health knowledge and information subsystem included the variables *own health research* and *knowledge about CKD and management*. It captured how health information influenced patients’ *motivation* and willingness to *engage with clinical care* and self-management, and how it was impacted by *doctor trustworthiness* and *community support*. Participants discussed how understanding of CKD among friends and family impacted their own knowledge and management of CKD. They discussed the need for a trusting, long-term relationship with a doctor to improve access to information, combined with access to reliable resources. The need for CKD knowledge was especially pronounced for patients who primarily managed another chronic condition, or had not known about their CKD diagnosis prior to the study.

The social support and responsibilities subsystem contained *community support* and *having time. Community support* could enable health behaviors directly, through making it easier to follow a CKD appropriate diet, and indirectly, by decreasing caretaking responsibilities and freeing up *time* for CKD management. Various types of community support needs included emotional support, practical support relating to caretaking and other time-consuming family and community responsibilities, as well as informational support. Patients discussed how lack of awareness about CKD, as well as cultural and societal values in their community, made it challenging to find support in managing their conditions. They mentioned a desire to participate in support groups for CKD patients.

Finally, the socioeconomic resources subsystem represented relationships between *having a job, money or income* and access to *insurance coverage*, and their impact on *engaging with* and *receiving clinical care. Money* and *insurance* impacted *engagement with clinical care*, representing the willingness or hesitancy patients might feel to initiate *engagement with clinical care* based on their ability to pay for it, and *receiving clinical care*, representing the impact of *money* and *insurance* to get continued and appropriate care. *Having a job* improved the ability to engage in health-promoting activities through having *money or income*, but it also decreased this via *having time* as a mediator. Participants discussed the political and structural aspects of these variables, including the need for universal health coverage, the for-profit nature of the U.S. healthcare system, and anxiety related to potential cuts to Medicare or Medicaid insurance.

While constructing the CLD, participants expressed that they found the experience deeply meaningful, and felt heard and supported. Workshop participants provided emotional and educational support to each other, exchanging information, advice, and encouragement. They emphasized the importance of self-advocacy for their needs with physicians, arguing that “nobody else is going to do it for you,” shared their stories of life after receiving a transplant, and celebrated each others’ milestones. During final reflections, participants noted that “[they] have a group of passionate individuals” and that “if [they] continue to mobilize, [they] could move change,” and indicated that they “will participate in other group activities.” Finally, participants demonstrated a sense of ownership over the CLD created, requesting that it be shared with them.

## 5. Discussion

Our CBSD study with CKD patients generated a formal representation of SDOH impacting patients with CKD and the relationships between those drivers, in the form of a CLD. To our knowledge, it is the first CBSD study with CKD patients. The variables in the final CLD spanned five subsystems, which broadly corresponded to four of the five SDOH domains identified by the Healthy People 2030 report (Pronk et al., 2021). Variables included are primarily intermediary determinants, impacting patients and their communities directly (Solar and Irwin, 2010). However, during focus groups and the workshop, especially during the final *dot exercise* for identifying intervention needs, patients discussed how additional structural determinants impact variables included in the CLD.

The resulting CLD can inform the development of clinical ML algorithms for CKD. For instance, it can support algorithm specification, particularly through incorporating variables in the graph as predictive covariates. For clinical algorithms that use only clinical and demographic variables, this enables the addition of patient-informed SDOH variables, with the potential for improving interpretation, performance, generalizability, and transportability (Foryciarz et al., 2025). Existing data sources, including data in the electronic health record and area-level social indices, may capture some variables in the graph. For variables that cannot be easily operationalized using existing data, this causal graph could inform the choice of proxy variables. Since the use of imperfect proxies derived from incorrect causal assumptions, such as race or health care costs, can reinforce inequities (Obermeyer et al., 2019; Vyas et al., 2020), the use of a system-level causal graph can promote an appropriate choice of proxies and lead to more fair and equitable decisions.

This graph can inform institutional efforts to collect SDOH data during clinical encounters, especially for variables that are currently unmeasured. Together with patient-identified needs discussed during the workshop (including improving access to information, nutrition support, social services and care coordination), the CLD can also serve as a starting point for the development of technological and non-technological interventions. Technical extensions to this work could consider how to adapt CLDs to generate causal directed acyclic graphs, or adapt GMB to directly generate causal directed acyclic graphs.

Beyond the CLD generated, the study accomplished several community- and education-related goals of CBSD, creating a space for emotional support for participants and serving a dual educational purpose for learning about CKD and system dynamics (Hovmand, 2014). The study allowed newly diagnosed patients to learn what life with more advanced CKD can look like, and learn about available resources. As evidenced in the final workshop reflections, the study mobilized participants to feel motivated to take action based on workshop insights, led to a sense of ownership of the final research product, and built community capacity and desire for further involvement in CBSD and research. Finally, it helped participants recognize the structural aspects of systems that influence their individual experiences, reframing their narratives about the root causes of situations (Hovmand, 2014).

The set of variables and connections identified by our study may have been impacted by our study setting and patient population. In particular, it reflects experiences of women with CKD with access to health insurance, and may exclude gender-specific perspectives impacting men with CKD and those without access to care. The focus on a single health care system facilitated study design and recruitment feasibility, and provided patients with a shared context, but may have additionally limited the set of identified variables.

More broadly, this work demonstrates the feasibility of applying participatory methodology to generate formal representations of complex causal theories that can inform the development of ML algorithms. By directly incorporating knowledge from multiple individuals who are directly experiencing social conditions impacting health, this approach is particularly conducive to generating explicit causal graphs including SDOH and their complex interactions. Utilizing this methodology can further improve algorithmic design and mitigate algorithmic bias for health applications.

## Data Availability

We use data collected from the patient screening survey and questionnaire, as well as notes and recordings generated during focus groups and workshops. The data, beyond summaries reported in the manuscript, are not available to other researchers for privacy reasons.

## Acknowledgments

We thank the Stanford Participant Engagement Program for their invaluable support in recruiting our study participants. Mock workshop participants— Oana Enache, Carter Nakamoto, Weston Hughes, and I. Elizabeth Kumar—provided crucial insight for our workshop development process. We are grateful to Paula Fleisher for insightful comments and discussion. This research was supported by National Institutes of Health Director’s Pioneer Award DP1LM014278.

## Appendix A. Group Model Building Sessions

**Table 1:**
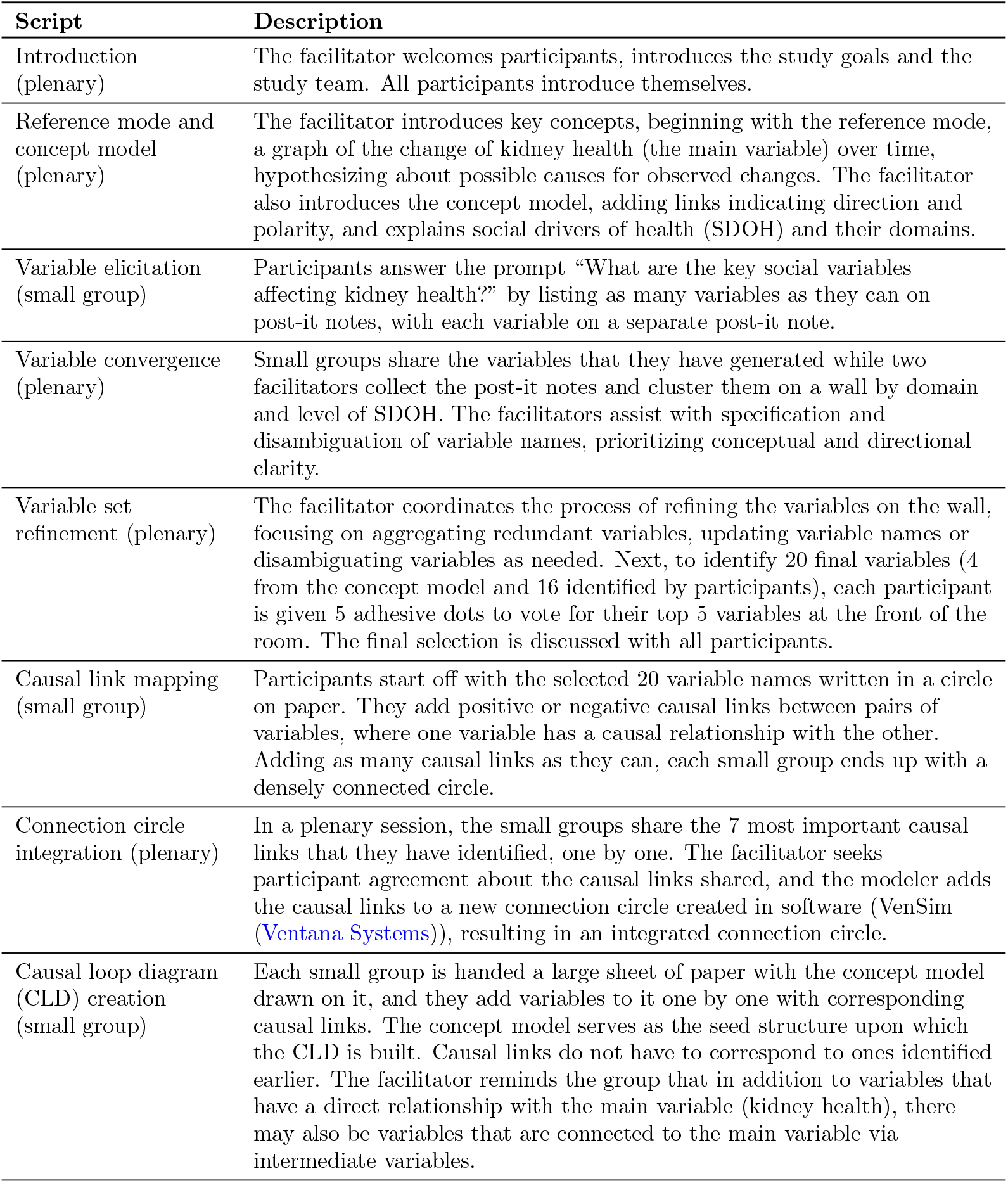
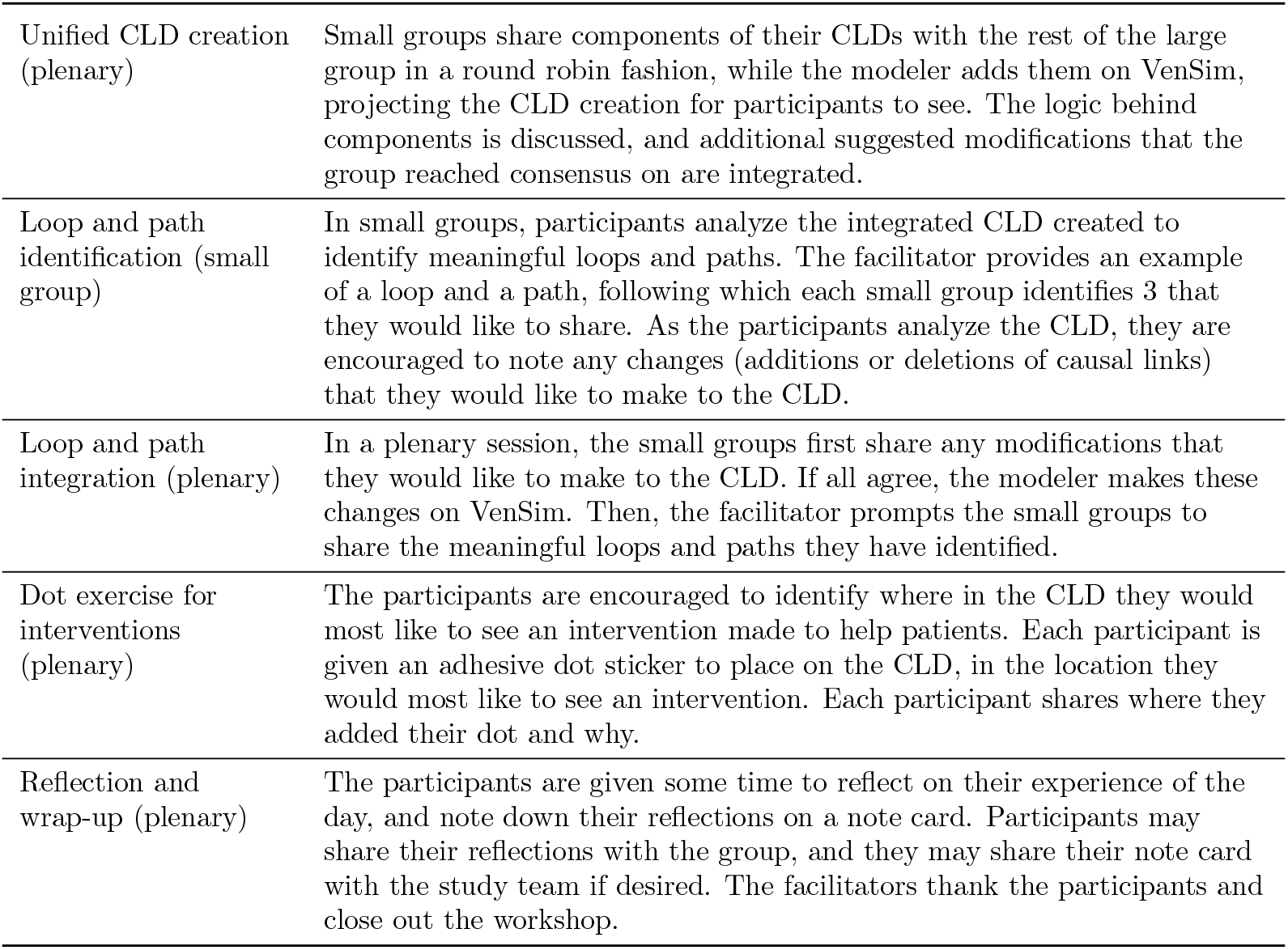
Description of group model building sessions.

## Appendix B. Focus Group Themes

The main themes that emerged in the focus group discussions were:

1. Nutrition & Diet
  a. Emphasis on nutrition and diet, tied to disease management and personal responsibility.
  b. Challenges around food choices and access to dietary guidance.
  c. Attention to water and salt intake.
2. Trust & Communication with Clinicians
  a. Positive and negative experiences with doctors.
  b. Issues of trust, feeling heard, personalized care, and shared decision-making (e.g. weighing benefits versus side effects of medications).
  c. Importance of seeing doctors in person, developing a trusting relationship over time, forming a partnership, switching physicians when necessary.
  d. Advocacy, including being your own advocate or having an advocate for proper care.
3. Comorbidities & Complex Health Management
  a. Multiple chronic conditions.
  b. Management of multiple diagnoses and medications.
  c. Health literacy, including discussions of trends in specific lab values over time.
  d. Frequent monitoring with telehealth support.
4. Transportation, Time & Systemic Barriers
  a. Time and logistical challenges, including time-consuming transportation, appointments, and messaging with care teams.
  b. Burdens of care navigation and “being a career patient.”
5. Work & Life Responsibilities
  a. Impacts of employment and caretaking on chronic kidney disease (CKD) management.
  b. Balancing work, caregiving, and healthcare.
  c. Need for work accommodations for doctor’s appointments.
6. Knowledge Gaps & CKD Awareness
  a. Variation in knowledge about CKD diagnosis, from lack of awareness about having CKD to being well informed.
  b. Need for early education, awareness, and access to resources at all stages.
  c. Informational resources from doctors and online sources, to understand the course of the disease and its management.
7. Emotional & Mental Health
  a. Stress, stigma, and fatigue.
  b. Emotional burden of living with a chronic condition, including resentment of the condition and feeling a lack of control.
  c. Need to “take control of your life.”
8. Social Support
  a. Family history, support systems, and communication with partners or relatives.
  b. Religion as a source of support.
  c. Social support as both a facilitator and a barrier.
  d. Social aspects of navigating nutrition, including challenges in dining out and having to explain.
9. Inequities: Race, Resources & Weight Stigma
  a. Racism, weight stigma, and inequitable access to resources and care.

## Appendix C. Original Causal Loop Diagram

**Figure 3.**
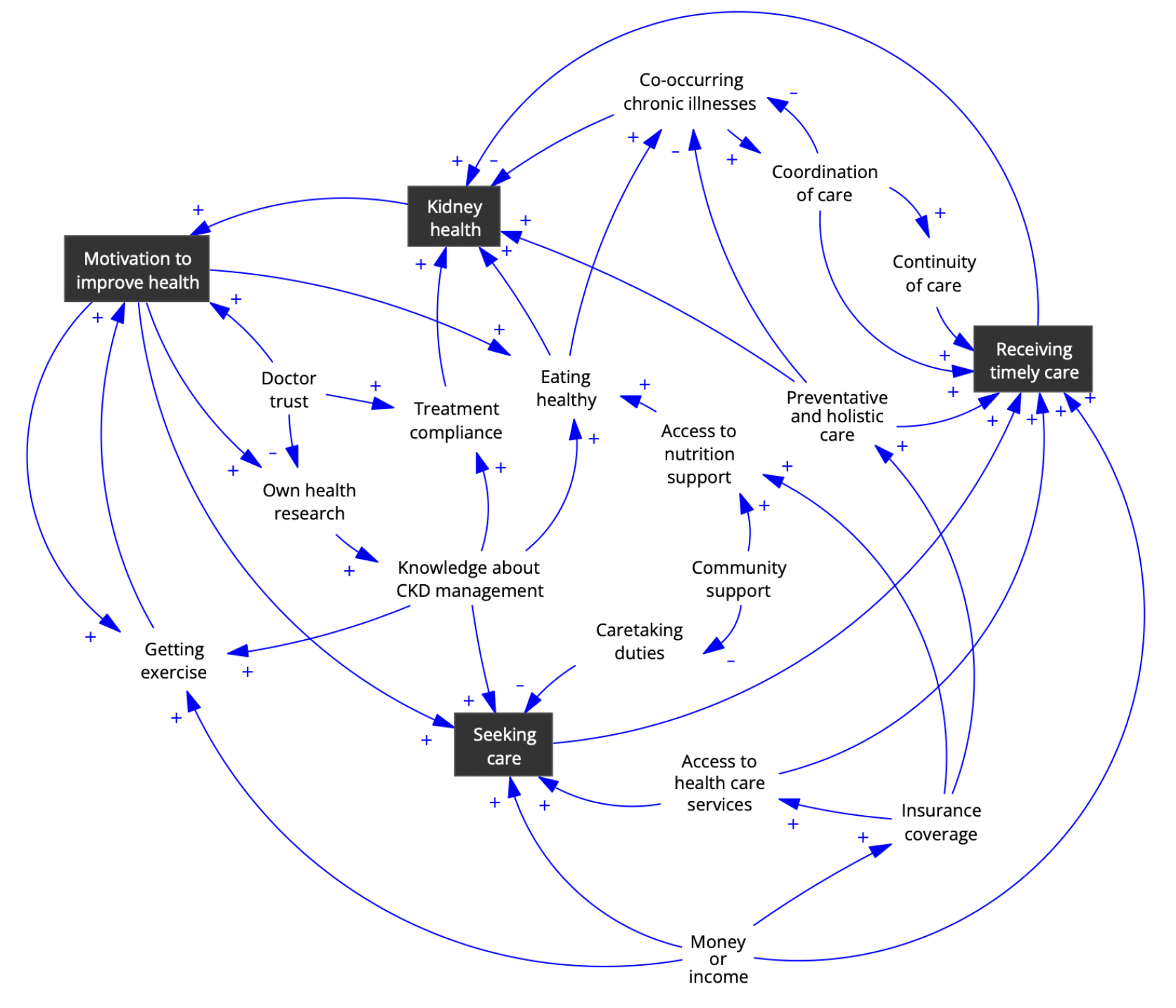
Original causal loop diagram created by participants during the workshop, prior to refinement by the modeling team. The variables in black boxes indicate those that were in the concept model.

## Appendix D. Recruitment

**Figure 4.**
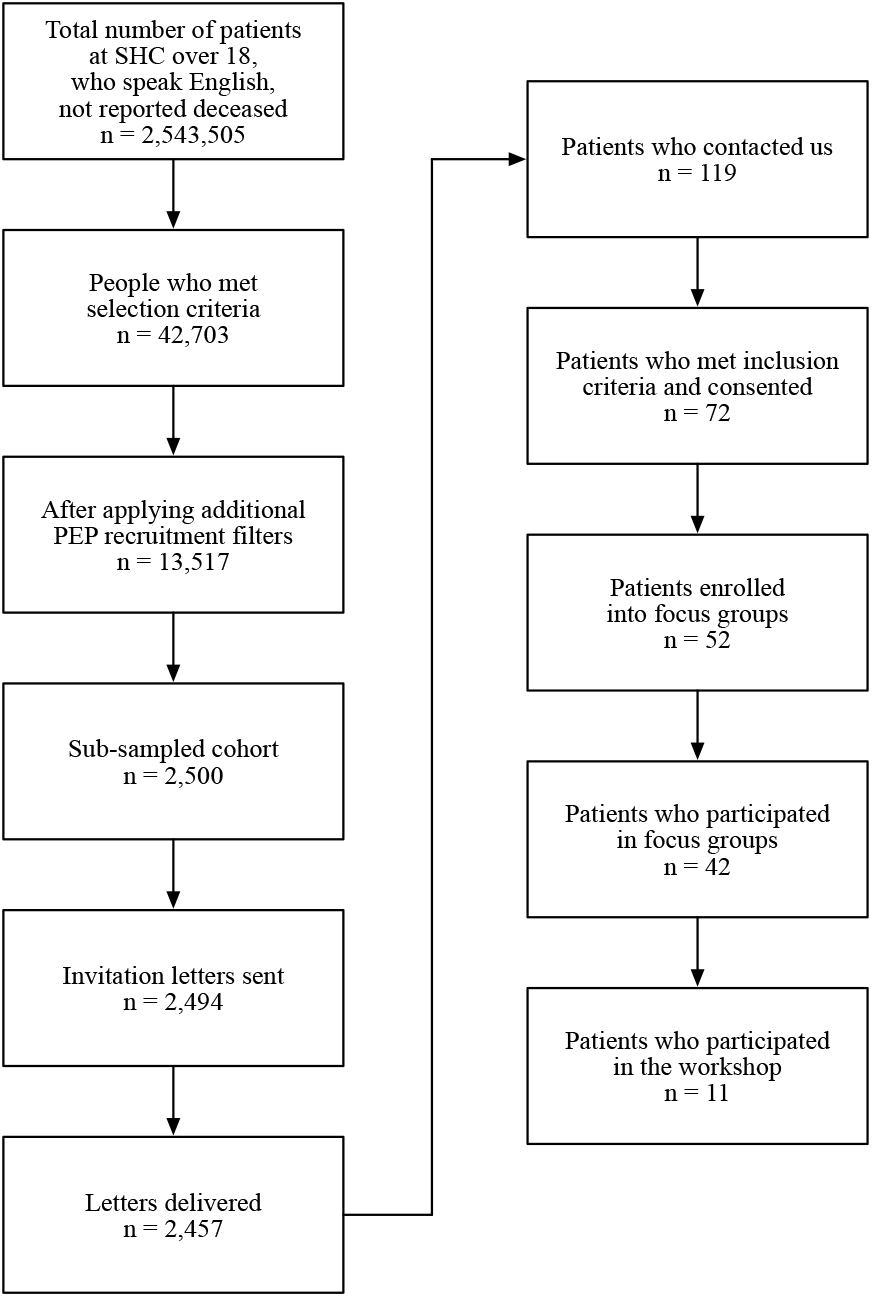
Recruitment flowchart.

