## Supplement for "Causal modeling of chronic kidney disease in a participatory framework for informing the inclusion of social drivers in health algorithms"

### Supplement 1: Focus Group Guide

The below focus group guide is included in its original form, unaltered from the time of conducting the focus groups. The only edit involves adding the table caption.

**1. Preparation (15 min)**

- Participant folders
- Compensations
- Pens
- Digital recorders
- Speaker order sheets
- Note pads

**2. Participant arrival & greeting (10 minutes)**

As patients arrive, both the facilitator and staff will greet and seat the patients, briefly introduce themselves.

**3. Informed consent + questionnaire (20 minutes)**

- Introduce to each participant study aims and procedures and give instructions to sign the consent form.
- When the consent forms are signed, provide the demographic questionnaire and let participants fill it out by themselves.

**4. Introduction + house rules (10 minutes)**

After ALL participants have arrived and completed the informed consent, the facilitator can start the introduction.

- ***WELCOME***
  - o *Welcome everyone! My name is______. We are from the Department of Health Policy at Stanford University School of Medicine, and we are studying specific ways in which social factors (e.g. housing, transportation, adverse experiences, access barriers, direct discrimination) impact patients' ability to manage chronic kidney disease (CKD). We will talk about what this is today in more detail.*

- ***STUDY GOALS***
  - o *The goal today is to better understand your experience with managing chronic kidney disease, and social determinants that have contributed to this experience. Your input will help us learn more about the best way to*

*engage community members in these efforts, and to identify potential needs for additional resources and programs addressing those social factors.*

- *In order to understand that, we will step back to define what social determinants of health are as well as how it might relate to you during the focus group. Don't worry if you have never heard the term or don't understand what it means because that information is also important for us to know and provides a starting point.*

- ***STUDY TEAM INTRODUCTIONS***
  - Now I would like to introduce our team.
  - [TEAM INTRODUCE THEMSELVES BRIEFLY]

- ***HOUSE RULES***
  - Before we begin, I have a few reminders for all of us. First, whatever we talk about today is anonymous, which means people will not find out what you said, but we will summarize what we talk about because it is a research study, which may include direct quotations.

  - It's important that we respect one another's privacy, and so I ask you to please not tell anyone outside of this group who was here or what was said.  Also, don't worry if you accidentally say someone's full name, because we will not write up that name on any paper.

  - You do not need to say your doctors' names when you talk about them. Also, don't worry if you accidentally say your doctor's name because we will not write up that name on any paper. And we will not share anything you say today with your doctors.

  - While we are asking your opinion and experiences with chronic kidney disease, you do not need to share details about your personal health if you do not want to. You can speak generally about your experiences or focus on other aspects of care related to chronic kidney disease.

  - So that we may have your full attention, it would be great if you could please turn off your cell phones or smart phones, or place them on silent. If you need to leave and step out to take a call briefly, that's perfectly fine.

  - There are no right or wrong answers. We are interested in your opinions and thoughts. Also, we are not looking for a consensus; so we don't expect you to agree with each other.

  - Lastly, we want to make sure that we are able to hear everyone's opinion. If you tend to talk a lot, I hope you won't mind if I interrupt so that others can voice their opinions.  If you are shy, I hope you'll try stepping out of your comfort zone and participate.

**Any Questions thus far?**

- ***PARTICIPANT INTRODUCTIONS***

    - Before moving into our group discussion, we would like each of you to state your first name and your favorite dessert (or sport or anything else).

#### 5. Group discussion (75 minutes)

**Note to Facilitators: Please try to cover all the prioritized questions (bolded) in each section and select the optional questions based on the flow of the conversations. Try to spend no more than 15 minutes on each section.**

**Supplemental Table S1: Focus Group Questions**

| | |
|---|---|
| Section 1. Experience with CKD diagnosis and care | |
| 1.a | **How would you describe CKD to someone who has not heard of it before?** |
| 1.b | **What aspects of CKD care have been most helpful in managing your symptoms and improving your health/halting progression?** |
| 1.c | **Has your CKD care been delayed at any point? If so, what contributed to the delay?** |
| 1.d | What's your relationship with your doctor managing your CKD? |
| 1.e | What types of services have you received from Stanford Health Care to help manage your CKD?<br>E.g. dietician consultations, visit scheduling support |
| 1.f | What kinds of conversations, if any, have you had with your physician about renal replacement therapy (dialysis/transplant) options? |
| 1.g | How does discrimination affect the care that patients with CKD get? |
| Section 2. Knowledge about CKD (education and health literacy) | |
| 2.a | **What would tell someone who wants to slow the progression of their CKD to do or not do?** |
| 2.b | **What non-medical resources do you think people need to slow the progression of CKD?**<br>E.g. financial resources, resources related to health |
| Section 3. Social and community context | |
| 3.a | **How has CKD affected your ability to fulfill your family and other social responsibilities?** |

| | |
|---|---|
| 3.b | **What type of support, if any, have you received from your immediate community (family, partner, friends, community groups) in managing CKD or other responsibilities which have been impacted by CKD?** |
| 3.c | Which of those were most significant and why? |
| 3.d | Have your family responsibilities or other relationships made it harder for you to get care? |
| 3.e | How has CKD impacted your family and other social relationships? |
| 3.f | How has the CKD diagnosis affected your stress levels? |
| 3.g | Have you received support related to your CKD or its consequences from any organizations? |
| Section 4. Neighborhood, built environment, transportation | |
| 4.a | **What makes it easy or hard for you to eat healthy?** |
| 4.b | What kind of barriers do you face in accessing healthy food? |
| 4.d | **What makes it easy or hard for you to be physically active?** |
| 4.e | Is it safe to go on walks in your neighborhood? Are there sidewalks and marked street crossing? |
| 4.f | Do you have access to recreational areas (e.g. parks, trails) nearby? |
| 4.g | **How easy is it for you to get to/from your medical appointments?** |
| 4.h | How do you get to/from your medical appointments? (e.g. bus, own car, other person's car, uber/lyft/taxi) |
| 4.i | What else about your neighborhood has been significant for your health? |
| Section 5. Economic stability | |
| 5.a | **How has CKD impacted your financial stability, job or housing?** |
| 5.b | What costs, other than medical costs, have you incurred? |
| 5.c | **How do you think the experience of CKD is different for more vs less affluent patients?** |
| 5.d | **Has your medical insurance been sufficient to cover the type of CKD care you need?** |
| 5.e | Have you been able to take advantage of sick leave or childcare to get medical care you need? |
| Section 6. Needs and additional questions | |
| 6.a | What additional resources do you think would be helpful for you, your provider or caretaker in managing your disease? |
| 6.b | What additional resources or programs do you think you and other patients could benefit from? |
| 6.c | How has pollution affected your symptoms or ability to get care? |

#### 6. Final comments & wrap up (10 minutes)

- Review the purpose of the focus group and see if anything has been missed or any other comments remain unvoiced.

- Summarize what you have learned with confirmation of the value of the participants' contributions.
- Express appreciation and dismiss in a timely manner.

### Supplement 2: Participant Questionnaire

Participants who enrolled in our study, to participate in a focus group or in the workshop, completed an additional questionnaire to collect information pertaining to demographics, CKD and medical history, and social determinants. The questionnaire questions are as below. This questionnaire is included in its original form, unaltered from the time it was administered.

**Introduction**

Thank you for your interest in our study on social determinants of chronic kidney disease (CKD) at the Stanford Health Policy Data Science Lab. Please fill out this short form to provide additional information about yourself. You have the right to refuse to answer particular questions. Your answers will be shared with our research team.

**I. Demographic information**

Which gender do you most identify with?
[ ] Female
[ ] Male
[ ] Non-binary / Additional group: ____________
[ ] Prefer not to say

What ethnicity do you most identify with?
Select one option.
[ ] Hispanic or Latino
[ ] Not Hispanic or Latino
[ ] Don't know / prefer not to say

Which race do you most identify with?
Please select one option.
[ ] American Indian or Alaska Native
[ ] Asian
[ ] Black or African American
[ ] Native Hawaiian or Other Pacific Islander
[ ] White
[ ] More than one race
[ ] Don't know / prefer not to say

For more than one race, which race(s) do you most identify with?
Select at least two options.
[ ] American Indian or Alaska Native
[ ] Asian
[ ] Black or African American
[ ] Native Hawaiian or Other Pacific Islander

[ ] White
[ ] Don't know / prefer not to say
[ ] Other: _______________

#### II. CKD/Medical history:

Have you ever been diagnosed with chronic kidney disease in the past?
[ ] Yes
[ ] No
[ ] Don't know

What is your stage of chronic kidney disease?
[ ] Stage 1
[ ] Stage 2
[ ] Stage 3
[ ] Stage 4
[ ] Stage 5 (end stage renal disease/kidney failure)
[ ] Don't know

Are you currently receiving dialysis?
[ ] Yes
[ ] No
[ ] Don't know

Have you received a kidney transplant?
[ ] Yes
[ ] No
[ ] No, but on the waitlist
[ ] Don't know

#### III. Additional information / Other social determinants

What is the highest level of education that you have attained?
[ ] Grade school (8 years or less)
[ ] Some high school (9-11 years of school in total, did not finish)
[ ] High School diploma or equivalent (GED)
[ ] Vocational or training school after high school
[ ] Some college or Associate's degree or technical certification
[ ] Bachelor's degree / undergraduate degree
[ ] Some graduate or professional school after college graduation
[ ] Graduate (MS, PhD, etc) or professional degree (MD, JD, DDS, etc)
[ ] Prefer not to say

What is your current job status? Mark the one that best describes you. If more than one describes you, please mark.
[ ] Employed (full time or part-time)
[ ] Homemaker, raising children, care of others
[ ] Disabled, unable to work
[ ] Not working
[ ] Prefer not to say
[ ] Other ___________________________________

What is your marital status?
[ ] Married
[ ] Living with a partner (Domestic Partnership)
[ ] Single or never married
[ ] Divorced
[ ] Separated
[ ] Widowed
[ ] Prefer not to say

Which category best describes how you usually pay for your medical care?
[ ] Pre-paid private insurance (for example: Health Maintenance Organization, Kaiser Permanente, or other Group Health-type plan)
[ ] Other private insurance (for example: Blue Cross, Aetna, etc.)
[ ] MediCare or MediCal
[ ] Military or Veterans Administration-sponsored
[ ] No insurance
[ ] Don't know / prefer not to say

### Supplement 3: Participant-Facing Group Model Building Terminology

To ensure that complex group model building (GMB) concepts are intuitive and conceptually clear to participants, we identified simple participant-facing terminology to use in the workshop, instead of more complex GMB terminology. We created a dictionary mapping GMB terminology to participant-facing terminology, provided in Supplemental Table S2. Although we use GMB terminology in our manuscript, we use participant-facing terminology in Supplement 4 (Workshop Schedule) and Supplement 5 (Workshop Scripts), and in the workshop itself.

Supplemental Table S2: Dictionary for Participant-Facing Group Model Building Terminology

| **Group Model Building Term** | **Participant-Facing Term** |
|---|---|
| Causal loop diagram (CLD) | Graph |
| Node *or* variable | Factor |
| Causal link | Link |
| Link polarities | Type of link *or* link type |
| Positive link polarity | Positive link |
| Negative link polarity | Negative link |
| Feedback loop | Loop |
| Reference mode variable | Main factor |
| Reference mode | Factor over time |
| Concept model | Simple graph |
| Connection circle | Link circle |
| Structural variables | Community factors |
| Individual variables *or* intermediary variables | Individual factors |
| Social drivers of health | Social factors |

### Supplement 4: Workshop Schedule

The day-long workshop consisted of four 60- to 90-minute sessions, with thirteen activities split amongst the four sessions and interspersed with frequent breaks. The schedule of the workshop is provided in Supplemental Table S3. This schedule is included in its original form, unaltered from the time of conducting the workshop.

Supplemental Table S3: Workshop Schedule

| Time (m) | Duration (m) | Agenda | Script | Plenary or small group? |
|---|---|---|---|---|
| 9:00–9:20 | 20 min | Participant arrival and greeting, coffee | | |
| 9:20–9:50 | 30 min | Introduction and overview, participant introductions | | |
| 9:50–10:10 | 20 min | Session 1 | Factor over time + simple graph script | Plenary |
| 10:10–10:15 | 5 min | | Break | |
| 10:15–10:30 | 15 min | | Factor elicitation script | Small group |
| 10:30–11:00 | 30 min | | Factor convergence script | Plenary |
| 11:00–11:15 | 15 min | Coffee break | | |
| 11:15–11:40 | 25 min | Session 2 | Factor set refinement script | Plenary |
| 11:40–11:45 | 5 min | | Break | |
| 11:45–12:00 | 15 min | | Link mapping script | Small group |
| 12:00–12:15 | 15 min | | Link circle integration script | Plenary |
| 12:15–1:00 | 45 min | Lunch break | | |
| 1:00–1:30 | 30 min | Session 3 | Small-group graph creation script | Small group |
| 1:30–2:00 | 30 min | | Unified graph creation script | Plenary |

| | | | | |
|---|---|---|---|---|
| 2:00–2:15 | 15 min | | Break | |
| 2:15–2:45 | 30 min | | Loop and path identification script | Small group |
| 2:45–3:15 | 30 min | | Loop and path integration script | Plenary |
| 3:15–3:30 | 15 min | Coffee break | | |
| 3:30–4:00 | 30 min | Session 4 | Dot exercise for interventions script | Plenary |
| 4:00–4:30 | 30 min | | Reflection and wrap-up script | Plenary |
| | | End (with 30 min buffer) | | |

Adapted from Andersen and Richardson: Scripts for Group Model Building (1996)[1]

### Supplement 5: Workshop Scripts

The scripts we used for our workshop are listed below, and were modified based on scripts from Scriptapedia[2]. Each script includes a speaking guide (typically indicated in italics) to assist the facilitators. The below scripts are included in their original form, unaltered from the time of conducting the workshop; the only edits made include adding captions to the figures, removing the names and initials of the facilitators, fixing typos, and adding references. In a few instances, a brief explanatory note is added in square brackets.

#### Introduction and overview

**1. Participant arrival and greeting (20 min)**

- As patients arrive, both the facilitator and staff will greet and seat the patients, briefly introducing themselves.
- Introduce to each participant study aims and procedures and give instructions to sign the consent form for any participants which had not already signed one.

**2. Introduction and overview, participant introductions (40 min)**

After ALL participants have arrived and completed the informed consent, the facilitator can start the introduction.

***Facilitator 1:***

- ***WELCOME***
    - o *Welcome everyone! My name is______. We are from the Department of Health Policy at Stanford University School of Medicine, and we are studying specific ways in which social factors (e.g. housing, transportation, adverse experiences, access barriers, direct discrimination) impact patients' ability to manage chronic kidney disease (CKD). We will talk about what this is today in more detail.*

- ***STUDY GOALS***
    - o *The goal today is to better understand your experience with managing chronic kidney disease, and social factors that have contributed to this experience. Your input will help us learn more about the best way to engage community members in these efforts, and to identify potential needs for additional resources and programs addressing those social factors.*
    - o *In order to understand that, we will step back to define what social factors of health are as well as how it might relate to you during the workshop. Don't worry if you have never heard the term or don't understand what it means because that information is also important for us to know and provides a starting point.*

- ***STUDY TEAM INTRODUCTIONS***
    - **o** Now I would like to introduce our team.

- [TEAM INTRODUCE THEMSELVES BRIEFLY]

***Facilitator 3:***

- ***HOUSE RULES***
    - Before we begin, I have a few reminders for all of us. First, whatever we talk about today is anonymous, which means people will not find out what you said, but we will summarize what we talk about because it is a research study, which may include direct quotations.

    - It's important that we respect one another's privacy, and so I ask you to please not tell anyone outside of this group who was here or what was said. Also, don't worry if you accidentally say someone's full name, because we will not write up that name on any paper.

    - You do not need to say your doctors' names when you talk about them. Also, don't worry if you accidentally say your doctor's name because we will not write up that name on any paper. And we will not share anything you say today with your doctors.

    - While we are asking your opinion and experiences with chronic kidney disease, you do not need to share details about your personal health if you do not want to. You can speak generally about your experiences or focus on other aspects of care related to chronic kidney disease.

    - So that we may have your full attention, it would be great if you could please turn off your cell phones or smart phones, or place them on silent. If you need to leave and step out to take a call briefly, that's perfectly fine.

    - There are no right or wrong answers. We are interested in your opinions and thoughts. Also, we are not looking for a consensus; so we don't expect you to agree with each other.

    - Lastly, we want to make sure that we are able to hear everyone's opinion. If you tend to talk a lot, I hope you won't mind if I interrupt so that others can voice their opinions. If you are shy, I hope you'll try stepping out of your comfort zone and participate.

  **Any Questions thus far?**

- ***PARTICIPANT INTRODUCTIONS***
  Before moving into our workshop, we would like each of you to state your first name and your favorite dessert (or sport or anything else).

***Facilitator 2:***

- ***OVERVIEW***
    - The goal of today's workshop is to generate a visual representation of the various social factors that are relevant to patients with chronic kidney disease, and their connections, called a graph.

- We will be using a method called group model building, which is used for mapping health and other public policy problems together with participants who have direct, lived experience.
- You will be guided through a step-by-step process where you will be asked to think about social factors and their connections, and discuss them with the group.
- We will now step back and define some concepts that we'll be using today, including what social factors impacting health are, and how these may relate to you during our workshop today. Don't worry if you have never heard the term or don't understand what it means because that information is also important for us to know and provides a starting point.

### Session 1

#### Factor over time + simple graph script

**Time:** 9:50–10:10 am
**Duration:** 20 min
**Format:** Plenary

**Materials:**
- Large sheets of paper (also called sticky easel pads)
- Large sheet of paper with 4 factors (for the simple graph) written on it in a circle
- Markers
- Projector

**Roles:**
- Facilitator: presents first portion of script (factor over time) to the entire group
- Modeler: serves as facilitator for the second portion of the script (simple graph), presenting this to the entire group
- Recorder: serves as facilitator for the third portion of the script (social factors), presenting this to the entire group; records group questions that arise

**Inputs:**
- Factor over time – drawn on large sheet of paper by facilitator
- Simple graph of 4 factors – drawn on large sheet of paper by facilitator

**Outputs:**
- Modified simple graph (modified with participant involvement)
- Familiarity with the main factor, and understanding that the main factor is dynamic (changing over time)
- Familiarity with a simple graph of 4 factors (including the main factor) and causal links

**Vocabulary: [terminology used in this script: other GMB terminology]**

- Factor: variable
- Factor over time: reference mode
- Simple graph: concept model
- Social factors: social drivers of health

**Steps:**

1. The facilitator introduces the main factor (of kidney health) and draws two factor over time trends, as shown below. The trends are drawn on large sheets of paper stuck on the wall. In the first, the main factor remains approximately constant over time, as shown by the blue line. In the second, shown by the orange line, the main factor shows a sharp decline and then plateaus.

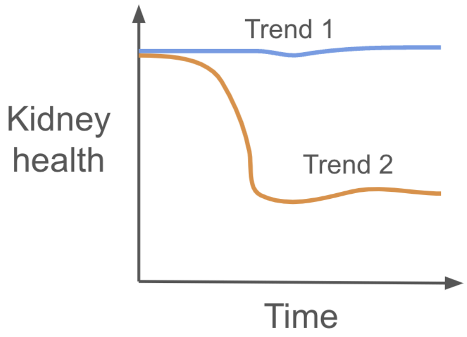

Supplemental Figure S1: Main factor over time.

2. The facilitator asks participants what the two trends represent, particularly what may have occurred at the point of decline and plateauing for the second trend line. Through this, the facilitator emphasizes that the main factor (kidney health) is a dynamic factor, changing over time.
3. The facilitator introduces that throughout the workshop, we will examine the factors that impact the dynamic behavior of the main factor. A facilitator handoff occurs here, transitioning from one facilitator to another, since all three facilitators will participate in this first script (the main facilitator, the modeler, and the recorder).
4. Facilitator introduces the simple graph (as below) by drawing it on a large sheet of paper stuck on the wall, mentioning that the additional factors cause or influence changes in the dynamic behavior of the main factor, either directly or through a causal chain. The facilitator introduces the links between these factors, and the type of link (positive link or negative link).

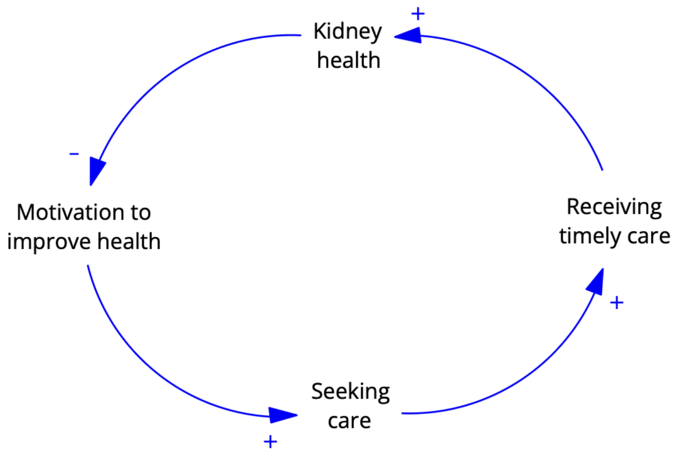

Supplemental Figure S2: Simple graph introduced by facilitator.

5. Facilitator asks participants whether they have modifications to suggest to the links between the factors in the simple graph. Facilitator asks participants whether they have additional factors to suggest that may influence any of the existing factors. Facilitator calls on 3-4 participants to share potential improvements, while adding them to the graph on the sheet of paper.

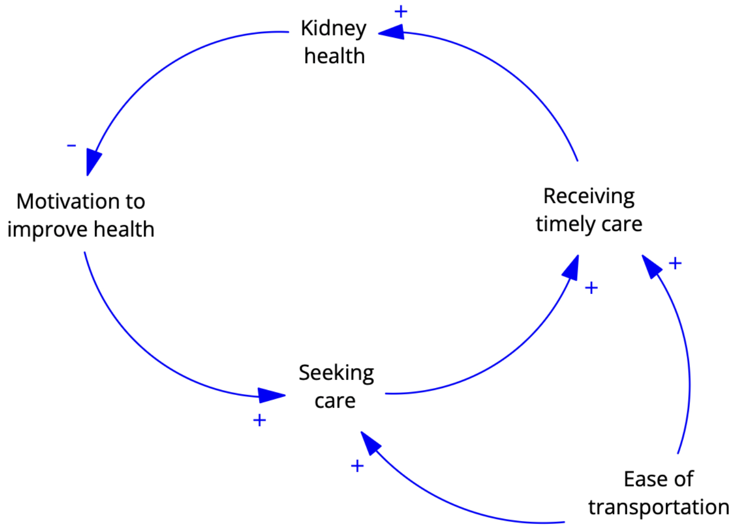

Supplemental Figure S3: Simple graph with potential added factor.

6. The facilitator shares that the group will be creating a larger, more comprehensive version of such a graph in the following sessions. A facilitator handoff occurs here, transitioning to the third member of the core modeling team who will facilitate.
7. The facilitator reiterates what factors are, and describes social factors and the domains of social factors. We will be focusing on social factors from all of the domains in the following sessions. The facilitator also mentions that social factors may be at the individual or community level, and provides an example of each.

***FACILITATOR 1 -***

*"The goal of our workshop today is to understand what factors contribute to changes in kidney function over time. To get us started, let's consider this chart. The direction of this arrow (point to y axis) indicates kidney function, with a perfectly healthy kidney on top, and a failed kidney at the bottom. The direction of this arrow (x-axis) is time, starting at birth.*

*Let's consider these two patients and their kidney health over time. This is the first patient (draw green line) and this is the second patient (draw red line).*

*[Image 1 - kidney health over time]*

*What can you tell me about these patients?*

- *Which one of them do you think might have CKD?*
- *What else do you notice?*
- *What may have happened here? (show decline)*
- *What about here? (show leveling out)*

*[Patients respond]*

*Thank you, great. This is exactly what we are going to be looking for today.*

***FACILITATOR 2 -***

*We will start by identifying <u>factors</u> that may impact our main factor (kidney health), and then thinking about <u>links</u> between them. Kidney health impacts and is impacted by a range of factors at different points in time, both directly and indirectly. A factor can be any "thing", and let's see some factors to better understand what factors are.*

*For example, [put up graph with just factors and no links between them], let's consider our main factor of kidney health, and these 3 additional factors that may influence kidney health:*

- *Motivation to improve health*
- *Seeking care*
- *Receiving timely care*

*Now, let's consider how these factors may be connected to or linked to each other. A worsening kidney health may make someone more motivated to improve their health and seek care, which could mean they are able to receive timely care. This will in turn improve their kidney health.*

*Here is how we could represent it in a graph. [Reads out the first link and uses this link to explain link direction and link type – whether the link is a plus or minus link].*

- *Being more motivated → more likely to seek care, **it's a positive relationship, mark +***
- *Seeking care → more likely to receive timely care, **it's a positive relationship, mark +***
- *Receiving care → improvement in kidney health, **it's a positive relationship, mark +***

- *Worse kidney health → more motivated to improve health,* ***it's a negative relationship, mark -***
    - *When one goes up, the other goes down, and we'll discuss this more later*
    - *[Can ask the participants what type of link they think this may be]*

*The direction of the links indicates the direction of influence between factors. A link begins at one factor and ends at another, indicating that the starting factor influences the ending factor. Links are also of two types: indicated by a plus or minus sign at the arrow of the link. [Refer to this as a plus link versus a minus link, or a positive link versus a negative link.] Plus links indicate that the factors change in the same direction, meaning that if the factor at the start of the arrow increases, the factor at the tip of the arrow also increases. If the factor at the start of the arrow decreases, the factor at the tip of the arrow also decreases. [Show thumbs up and down for this.] Negative links indicate that the factors change in opposite directions, so if the factor at the head of the arrow increases, the factor at the tip would decrease, and vice-versa. Just to be clear, plus and minus doesn't mean good or bad, it just shows whether the two factors in the link change in the same direction, or change in opposite directions.*

*Do the links to this graph make sense to everyone? Are there any changes you would recommend? [Asks 1-2 participants who raise their hands to share potential changes, asks the rest of the group if the changes make sense, and adds them to the graph if consensus is reached]*

*Now, let's think about how we could add another factor to this graph, and capture how it influences these factors that are already in our graph. Something that has come up during focus groups is the importance of transportation and challenges associated with it. So, let's add a factor called 'Ease of transportation' to our graph. [Writes ease of transportation in the lower right corner]*

*Are there any links you would like to add between 'Ease of transportation' and any other factor on the graph? [Waits for 1-2 suggestions from participants. If the group agrees on the suggestion, add the link to the graph.]*

*[If no suggestions are provided] What about if we consider the factor of 'Seeking care' – is there a link between 'Seeking care' and 'Ease of transportation'? [Wait for suggestions from participants, add suggestions to the graph]*

*Great, we have now created a simple graph, showing some factors impacting kidney health and the links or connections between these factors. What we'll be doing over the course of the day today is create a much larger, more comprehensive version of such a graph.*

***FACILITATOR 3 -***
*In particular, we will be interested in social factors. Those are "conditions in the environments, such as where people are born, live, learn, work, play, worship, and age that affect a wide range of health outcomes." They are "nonmedical factors that can make it harder for you to be healthy*

*and get the medical care and support you need." Some examples of social factors that have come up during focus groups include*

- *[TODO]*

*[Display domains of social factors on the projector.]:*

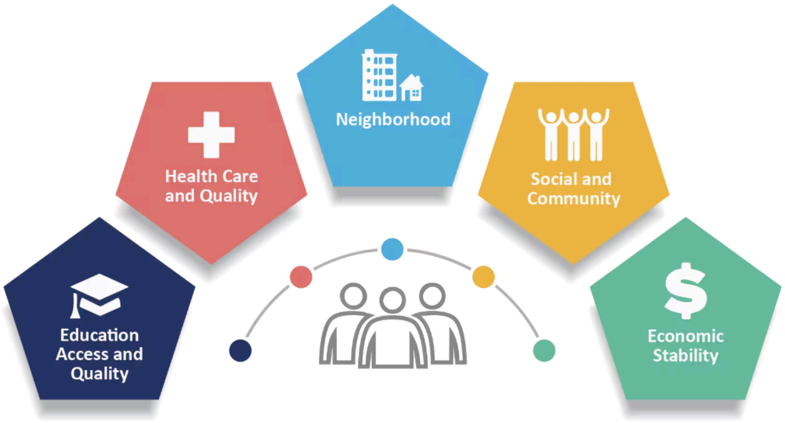

Supplemental Figure S4: Social factors domains introduced to participants.[3]

- *Remove "social determinants of health", all of us → social factors*

**References:**

1. This script is modified from *Scriptapedia: Presenting the Reference Mode*[4]
2. *Scriptapedia: Concept Model*[5] (this concept model script is for a stock and flow diagram, not a causal loop diagram, but has roughly the same structure)
3. (for text)
    a. *National Kidney Foundation: Social Determinants of Health and Chronic Kidney Disease*[6]
    b. (image #1): *Centers for Disease Control and Prevention: Social Determinants of Health*[3]

#### Break

**Time:** 10:10–10:15 am
**Duration:** 5 min

#### Factor elicitation script

**Time:** 10:15–10:30 am
**Duration:** 15 min
**Format:** Small group

**Materials:**

- Post it notes
- Markers
- Projector

**Roles:**
- Modeler: introduces / facilitates this script
- Process coach: supports small groups groups who may require assistance

**Inputs:**
- NA

**Outputs:**
- List of factors per small group (no limit on the number of factors)

**Vocabulary: [terminology used in this script: other GMB terminology]**
- Factor: variable
- Social factors: social drivers of health

**Steps:**
1. The facilitator divides the large group into small groups of ~4 people per small group. Each small group is given post-it notes and markers.
2. The facilitator projects a slide with a task focusing question: “What are the key social factors affecting kidney health?”
3. The facilitator asks each small group to write as many problem-related factors as they can on the post-it notes, such that each factor is listed on its own post-it note. Factors from the simple graph or from the focus groups are provided as examples. The small groups are given 10 minutes to generate their set of factors.
4. When there is 1 minute left to go, the facilitator provides a 1-minute warning, and asks the small groups to select one person in each small group to share the list of factors with the larger group (in the next script).

***FACILITATOR 2 -***

*The goal of this session is to identify factors impacting kidney function. We'll work in groups of 3-4. You'll be asked to answer the question* ***“What are the key (social) factors affecting kidney health?****” (displayed on the projector). For the next 10 minutes, think of as many factors as possible, and write each on a separate post-it note. You may talk with each other in the group as you are generating those. We will then share them with the entire group.*

*[time to divide into groups, each person in the group is given a block of post-it notes and a marker - make sure they're all the same color.]*

*[at the 9 minute mark] You have 1 minute left. We will ask each group to identify one person who will share the factors with the rest of the group, in the order of importance.*

**References:**

1. This script is modified from *Scriptapedia: Variable Elicitation*[7]

#### Factor convergence script

**Time:** 10:30–11:00 am
**Duration:** 30 min
**Format:** Plenary

**Materials:**

- Large sheets of paper
- Post it notes
- Tape
- Markers (of multiple colors)

**Roles:**

- Facilitator: Facilitates session; collects factors from small groups and clusters them
- Modeler: Assists facilitator in collecting post it notes and clustering (as a wallbuilder)
- Recorder: Captures the products of the process photographically and in the form of notes

**Inputs:**

- List of factors per small group (no limit on the number of factors)

**Outputs:**

- Clustered set of factors collected from all the small groups

**Vocabulary: [terminology used in this script: other GMB terminology]**

- Factor: variable
- Simple graph: concept model
- Individual factor: individual variables
- Community factor: Structural variables

**Steps:**

1. The facilitator draws a human with two kidneys on a large sheet of paper taped on a wall. The facilitator states that the group is going to add all the factors generated to the large sheet of paper, and begins by adding the 4 factors from the simple graph to the sheet.
2. The facilitator requests the small groups to share their factors one at a time in a round-robin fashion, in the order of importance – that is, one person from each small group shares the groups next most important factor, until all factors from all small groups have been shared.
    a. When a factor name is vague or otherwise ambiguous, the facilitator asks for a brief description or definition of the factor, ensuring that the factor is specific and has a clear direction (by direction, ensuring that it is clear what it means for the factor to increase).

    b. When necessary, the modeler may use a marker to modify the factor on the post-it note, or to split up the factor into multiple factors (on multiple post-it notes). Thus, there may be 1:1 or 1:many conversions at this stage. However, there will not be many:1 conversions, and it is alright to have multiple versions of the same factor generated.
    c. The conversions can either be done directly on the same post-it note (for 1:1 conversions) if possible, or on new post-it notes. During conversions, the old factor(s) and removed from the sheet of paper, and the new factor(s) are added to the sheet of paper.
3. The <u>facilitator and the modeler</u> tape each post-it note, as it is shared, on the large sheet of paper.
    a. This may be done by the modeler collecting the post-it note from the participant and suggesting a location on the wall which the facilitator may modify, or the two may resolve it through minor discussions.
    b. The post-it notes are clustered such that they form thematic clusters and are located at the appropriate level – with individual factors being placed closer to the human figure on the paper and community factors being placed further from the human.
4. Once all of the factors have been shared, the facilitator returns to the 4 factors from the simple graph and asks participants whether they would like to make any 1:1 changes (modifications to the factor name) for the 3 factors other than the main factor – ‘Motivation to improve health’, ‘Seeking care’, and ‘Receiving timely care’.
5. The facilitator reflects back the thematic clusters that have emerged, showing that the factors fit into 4-5 sub-domains. The facilitator circles each sub-domain with a colored marker. The facilitator also shares that the factors closest to the human are individual factors while the factors further from the human are community factors. The facilitator asks the participants for feedback, including asking questions such as “Does this resonate with you? Are there other themes you notice, or any factors you think should be moved to a different sub-domain? Should any factors be moved to a different level – from individual level to community level or vice-versa?”
6. The recorder captures the products of the wall building process photographically.

***FACILITATOR 1 -***
*[Draws human]*

*I will ask each group to share a single factor at a time, and we’ll keep going around until all factors are shared. I’ll put those factors up on the board.*

*[As I’m putting post-its on the board]*

- *[order]*
    - *Order (I don’t share this with participants, but that’s what I’ll be implicitly using):*
        - *Inner circle: biological factors*
        - *Middle circle: individual- or immediate community-level factors*
        - *Outer circle: structural factors*

    - *[inner "circle"] I'm going to put this one close to the person, since this factor impacts us <u>directly/individually</u>*
    - *[middle "circle"] I'll put this one a little further, since <u>it is impacted by and impacts our immediate community as well</u>*
- *[interpretation]*
    - *[same] I'll put these two together, since they relate to the same thing*
    - *[related] I'll put it close to …, since they seem related*
    - *[clarity] Could you describe this briefly?*

*[Once all have been shared] Now that all the factors have been shared, let's look back at them and think about some themes that are coming up.*

*[Read out names of all factors. Let's now group some of these factors together into themes. It seems that these all relate to the health system, do you agree? I'll put a green circle around them. These are all transportation. …*

*As I was placing them on the board, I was organizing them both in relation to each other - placing those with similar meanings close together - but also in relation to the individual in the middle. We can think of factors as existing on two levels:*
- *Inner circle: individual factors (biological, behavioral and social position factors: nutrition, stress levels, other diseases, nutrition, smoking, job type, income)*
- *Outer circle: community-level factors (i.e. healthcare access, family support)*

*Many factors which influence kidney health do it indirectly - community factors might impact individual factors, like patients' behavior, or the care they're able to receive, before kidney health is impacted. We're hoping to understand those relationships today.*

**References:**
1. This script is modified from *Scriptapedia: Variable Elicitation*[7]

##### Coffee break

**Time:** 11:00–11:15 am
**Duration:** 15 min

**Roles:**
- Facilitator and Modeler: Discuss a strategy for distillation/refinement of factors in the next script.

#### Session 2

##### Factor set refinement script

**Time:** 11:15–11:40 am

**Duration:** 25 min
**Format:** Plenary

**Materials:**
- Post it notes
- Tape
- Markers

**Roles:**
- Facilitator: Facilitates session
- Modeler: Helps facilitator determine the top 16 factors
- Recorder: Captures the products of the process photographically and in the form of notes

**Inputs:**
- List of factors generated and clustered

**Outputs:**
- List of 20 final factors

**Vocabulary: [terminology used in this script: other GMB terminology]**
- Factor: variable
- Social factors: social drivers of health
- Individual factor: individual variables
- Community factor: Structural variables

**Steps:**
1. The facilitator states that in this session, the group will be refining the set of factors generated, and then will vote on a smaller set of factors to work with for the remainder of the workshop.
2. The facilitator then begins the process of refining the factors on the wall, focusing on aggregating redundant factors (many:1 conversions). In cases where 1:1 conversions of factor names or 1:many conversions to disambiguate factors are required, these may also be conducted, although most of these will have been completed while collecting the factors in the factor convergence script.
    a. When making conversions, the post it notes for the old factor(s) will be removed from the sheet of paper and the post it notes for the new factor(s) will be added.
3. Once all factors have been refined, the facilitator states that the group will together select 16 factors to work with for the remainder of the day, in addition to the 4 factors from the simple graph. The facilitator indicates the 4 factors from the simple graph by adding stars (using a marker) to the respective post-it notes.
4. The modeler hands each participant 5 adhesive dots, while the facilitator states that each participant can use their 5 adhesive dots to vote for their top 5 factors. The

participants are given 5 minutes to take turns coming to the front of the room and place their adhesive dots on 5 post-it notes each.

5. As the group finishes voting, the facilitator and modeler determine the top 16 factors, which the facilitator shares with the group. This set of 20 factors (including the 4 factors from the simple graph) will be used for the remainder of the workshop.
6. The recorder captures the products of the process photographically and in a word processor.

***FACILITATOR 1 -***

*As above.*

*[Once all are all board, reflect on themes that emerged, ask participants for feedback]*

*[**Refining factors**] Now, we have a large number of factors, and some of them overlap in meaning. Let's try to refine some of the terms. The goal is to*

1. *Clarify factors*
2. *Combine post-its which refer to the same factor*
3. *Later, we will reduce the number of factors so we have a set of about 20 to work with*

*We will keep the*

*There are three factors we put together here: "healthy food", "nutrition" and "cooking". Do you think these all relate to the same thing, or are they different? How about "eating healthy" and "cooking skills"?*

- Types of refining
    - 1→1 (clarify)
    - 1→many (disambiguate)
    - Many→fewer (reduce, clarify)
- Reduce in plenary (to get about ~15-20, can be flexible about how granular we want factors to be)

*[**Reducing factors**]*

*Our next step is to reduce the number of factors so we have a set of about 20 to work with. You will each be given 5 stickers, and we'll ask you to come up to the front and place them on the factors you'd like to keep.*

*We will keep the 4 factors from the single graph, which we'll mark now with stars - so no need to vote for those. We'll ask each group to come up one by one, staring with group 1 [point], so we're not too crowded.*

*[Once voting is complete] Ok, it looks like the factors that got the most votes are … [read them out]. We will not consider [read it out].*

*These are the factors that we will primarily be working with for the remainder of the day.*

*Now, we'll take a 5 minute break - feel free to step out of the room. We'll meet back here at [give time].*

**References:**

1. This script is modified from *Scriptapedia: Variable Elicitation*[7]

#### Break

**Time:** 11:40–11:45 am
**Duration:** 5 min

**Materials:**

- Printer
- A4 sheets of paper

**Roles:**

- Additional team member: creates and prints out link circle templates with the final list of factors identified.
    a. To prepare the link circle template, the team member uses a laptop to create a document with the final list of factors in a circle, except the main factor (kidney health). A gap is left for the main factor to be later added, as shown below. This document is printed out on A4 sheets of paper, with each link circle template spread out across 4 sheets of paper. The sets of 4 sheets of paper are taped together to make large link circle templates. Six such templates are prepared.

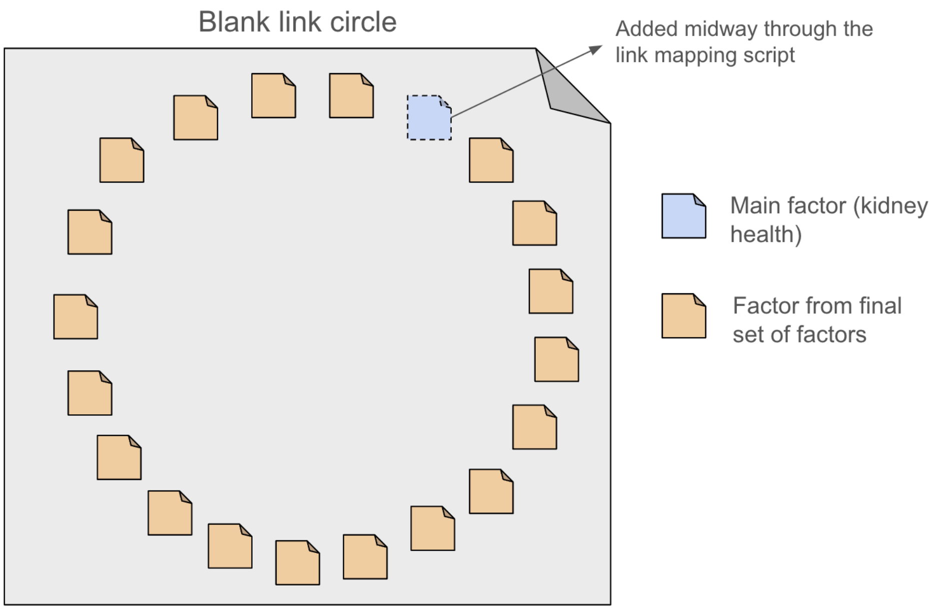

Supplemental Figure S5: Link circle template.

#### Link mapping script

**Time:** 11:45 am–12:00 pm
**Duration:** 15 min
**Format:** Small group

**Materials:**
- Markers
- Link circle templates

**Roles:**
- Modeler: introduces / facilitates this script
- Process coach: supports small groups groups who may require assistance

**Inputs:**
- One blank link circle template per small group + one for facilitation

**Outputs:**
- One filled in link circle (with links added) per small group

**Vocabulary: [terminology used in this script: other GMB terminology]**
- Factor: variable
- Main factor: reference mode variable / main variable
- Link: causal link
- Link type: link polarity
- Positive link: positive link polarity
- Negative link: negative link polarity
- Link circle: connection circle

**Steps:**
1. The facilitator instructs the large group to return to their small groups. Each small group is given a printed blank link circle. At this stage, the blank link circles do not have the main factor (kidney health) added, and instead have a blank spot left in the circle where the main factor (kidney health) will later be added.
2. The facilitator introduces the exercise by stating the goal of identifying the links between the factors that the group has identified. The facilitator states: "*We are going to draw a link circle. A link circle is a visual tool that can help us identify and understand problems and see the connections in a system. First, let me show you an example*."
3. The facilitator tells participants, "*We are going to start with a large circle with the factors we have come up with*." The facilitator hangs up a blank link circle on the wall. Next, the facilitator explains that the participants will then pick two factors that are connected and draw a link between them: an arrow pointing in the direction of influence. The arrow shows causality and it can indicate a positive or a negative link. The facilitator provides an example to the participants, by choosing two factors, reasoning about the direction of

influence between them, whether this direction of influence is a positive or negative link, and then drawing the link on the sheet of paper using a marker.

4. The facilitator says: *"Next, you will pick another set of factors that are connected and draw an arrow to show causality. As you continue to do this, you will have a web of connections between all your factors."*
5. The facilitator tell the participants that there are several points to keep in mind before starting:
    a. First, for a link that goes in both directions, two separate links should be drawn, one going in one direction and the other going in the other direction.
    b. Second, it may be easier to bend some of the lines to make them easier to follow, and that's OK.
    c. Third, links can be based on personal experiences.
    d. Fourth, this link circle is the overall or combined group picture of what may be happening in relation to kidney health. Some factors and links may be common to all communities. Other factors and links may be specific to only one community or group. So, they can add links even if they only apply to some people, not everyone.
    e. Fifth, it is ok if some factors have many links, and some can even have none.
    f. Finally, when adding a link, it may be helpful to think about if the link between the two factors being considered is a direct link, or whether it may be that a factor influences another through an intermediate third factor.
6. The facilitator tells participants that they have 7 minutes to work on this exercise in their small groups. As the small groups are working on this activity, the facilitator and the process coach walk around and support any groups that require assistance.
7. After the 7 mins are complete, the facilitator states that we are now going to add the main factor of kidney health to the circle and continue to add links, focusing now on links that relate directly to the main factor. The facilitator hands each small group a post-it note with the main factor to add to the empty spot in the circle. The facilitator tells participants that they have an additional 3 minutes to work on this exercise in their small groups. [Note added: We initially excluded the kidney health (main) variable in the small group link mapping, for the first 7 minutes, and then added it for the last 3 minutes. This was to encourage the identification of causal connections between any pair of variables and indirect links between variables and kidney health.]
8. When time is approaching the end, the facilitator reminds the small groups that one person in each small group will share their link circle with the larger group.

***FACILITATOR 2 -***
*In this session, we will identify connections between pairs of factors. We are going to start with a large circle with the factors we have come up with, and draw links between any two pairs of factors we believe are connected. First, let me show you an example. Let's pick two factors that we think are connected. Let's say [blank1] and [blank2]. What's the direction of the link here? Which factor is impacting which?*

*[Participants answer]*

*What about the type of link, is it positive or negative? When [blank1] increases, what happens to [blank2]?*

*[draw link]*

*We'll ask you to work in groups again, using a copy of the circle we have created for you. You will pick another pair of factors that are connected and draw an arrow to show they are linked. And you will add whether the link is a plus link or a minus link. As you continue to do this, you will have a web of connections between all your factors. We'll end up with a link circle.*

*A couple of points to keep in mind before starting:*

1. *First, for a link that goes in both directions, two separate lines should be drawn, one going in one direction and the other going in the other direction. That the arrow shows the direction of influence, or of causation. The arrow can represent something positive or negative.*
2. *Second, it may be easier to bend some of the lines to make them easier to follow, and that's OK.*
3. *Third, link can be based on personal experiences.*
4. *Fourth, this link circle is the overall or combined group picture of what may be happening in relation to kidney health. Some factors and connections may be common to all communities. Other factors and connections may be specific to only one community or group. So, they can add connections even if they only apply to some people, not everyone.*
5. *It is ok if some factors have many connections, and some can even have none.*

*We'll be walking around and happy to answer any questions. If it is unclear what an increase or decrease in a factor means, please bring it up. We'll have 10 minutes.*

*[After 7 minutes]*

*We are now going to add the main factor of kidney health to the connection and continue to add connections, focusing on connections that relate directly to the main factor. You'll have 3 more minutes.*

*[1-minute warning]. We'll ask one person from each group to share out links your group has identified with the larger group, one by one. Each group will share 5 links, so think about which ones feel important to you.*

**References:**

1. This script is modified from *Scriptapedia: Connection Circle*[8]

#### Link circle integration script

**Time:** 12:00–12:15 pm
**Duration:** 15 min
**Format:** Plenary

**Materials:**

- Blank link circle

**Roles:**

- Facilitator: Collects links from the small groups
- Modeler: Tapes up each small group's link circle on the wall, at the beginning of the activity. During the activity, the modeler projects a blank link circle on VenSim[9] and adds the links suggested as the groups share them.
- Recorder: Captures the products of the process photographically, for each small group's link circle and for the integrated link circle

**Inputs:**

- One filled in link circle per small group
- One blank link circle (on VenSim)

**Outputs:**

- One filled in link circle (on VenSim) integrating the small groups' link circles

**Vocabulary: [terminology used in this script: other GMB terminology]**

- Factor: variable
- Main factor: reference mode variable / main variable
- Link: causal link
- Link type: link polarity
- Positive link: positive link polarity
- Negative link: negative link polarity
- Link circle: connection circle

**Steps:**

1. The modeler projects on the screen a blank link circle on VenSim. The facilitator informs the group that we will now be integrating the link that each group has made into a shared understanding of the links between the factors.
2. The facilitator has the small groups share the links in their link circles in a round-robin fashion – that is, one person from each small group shares the next most important causal link from the groups link circle, until each small group has shared 7 links. As each link is shared, the facilitator prompts the participant sharing to provide a brief description of the link and the link type (positive or negative). As each link is shared, the modeler adds the link to the link circle on VenSim.
3. The facilitator ends the session by noting that the integrated link circle represents a combination of each group's links between factors.

4. The recorder captures the products of the process photographically.

***FACILITATOR 1 -***

*As before, we'll now ask you to share your links one at a time, describing the direction and the sign. [Facilitator 2] will add each link that's shared on the screen, so we can all see it. As you'll sharing, we'll ask you to briefly explain your links.*

*[As links are shared]*

- *Could you briefly explain this link?*
- *What's an example of when this connection would show up?*
- *And which is impacting which?*
- *When xxx increases, does xxx increase or decrease?*

*[When complete] The integrated connection circle represents a combination of each group's connections between factors.*

*Thank you everyone! We've all done so much work already. We will now go into lunch for an hour. When we return, we will analyze the connections we've identified, and start exploring a larger graph structure.*

**References:**

1. This script is modified from *The Systems Thinker: Learning About Connection Circles*,[10] and *Scriptapedia: Variable Elicitation*[7] (for going around the room, etc.)

#### Lunch break

**Time:** 12:15–1:00 pm
**Duration:** 45 min

**Materials:**

- Modeler's laptop with VenSim
- Facilitator's laptop with VenSim (as a backup)

**Roles:**

- Modeler: Runs script to generate spatially re-organized graph given the link circle as input, using GraphViz.[11] Generates a list of loops that exist in the link circle using VenSim.
- Facilitator, Modeler: Discuss the GraphViz figure and the loops generated via VenSim to understand some initial stories that may emerge in later stages of the workshop. Select a factor and link to add to the simple graph as an example, in the first stages of the next script. + select an example of a factor not directly connected to kidney health
- Additional team member: Creates 6 copies of the simple graph on large sheets of paper, via drawing with a marker.

### Session 3

#### Small-group graph creation script

**Time:** 1:00–1:30 pm
**Duration:** 30 min
**Format:** Small group

**Materials:**

- Large sheets of paper
- Markers
- Projector

**Roles:**

- Facilitator: introduces and facilitates this script
- Process coach: supports small groups groups who may require assistance

**Inputs:**

- Final set of factors
- Understanding of potential links between factors from link circle

**Outputs:**

- One graph per small group

**Vocabulary: [terminology used in this script: other GMB terminology]**

- Factor: variable
- Link: causal link
- Link type: link polarity
- Positive link: positive link polarity
- Negative link: negative link polarity
- Graph: causal loop diagram
- Simple graph: concept model

**Steps:**

1. The facilitator reviews the integrated link circle formed at the end of the previous script, and shares that the group will now use this understanding of links between factors to generate graphs in small groups.
2. The facilitator tapes up a large sheet of paper containing the simple graph, and projects a slide with a list of the final factors. The facilitator demonstrates adding a factor to the simple graph.
    a. This is done by first choosing a factor that is on the final list of factors but not yet added to the simple graph. This factor may contain a link to/from one of the existing factors in the integrated link circle.

   b. This factor is added to the simple graph along with its link to an existing factor in the simple graph, and the facilitator demonstrates adding any additional links that may exist from this added factor to the existing factors.
   c. The facilitator shares that we can continue adding all the factors from the final list of factors to the graph in this manner, adding all appropriate links as we proceed.
   d. The facilitator emphasizes that when factors are being added, they can be spatially located wherever is most convenient, which may often be closest to the other factors that they are linked to.
   e. The links in the integrated link circle are a suggestion, but additional links that were not identified in the link circle can be added to the graph, and all links from the link circle need not be added to the graph.
3. The facilitator instructs the group to return to their small group, and instructs them that each small group will generate a graph in this manner. Each small group is handed a large sheet of paper with the simple graph drawn on it.
   a. The facilitator suggests that it may be helpful for one person in a small group to make a suggestion to their small group, and then to check with other group members if they agree with the proposed addition. If someone disagrees, the group can discuss to try to determine what the group thinks the relationship should be. If a discussion goes on too long, the group can choose to temporarily ‘park’ this item and continue with another part of the graph.
   b. The facilitator reminds the group that in addition to factors that have a direct relationship with the main factor (kidney health), there may also be factors that are connected to the main factor via intermediate factors.
4. The facilitator tells participants that they have 15 minutes to work on this exercise in their small groups. As the small groups are working on this activity, the facilitator and the process coach walk around and support any groups that require assistance.
5. When there are 2 minutes remaining, the facilitator provides a warning, and reminds the small groups that one person in each small group will serve as the group speaker to share with the larger group.

***Facilitator 2:***
*[Display link circle]*

*Welcome back! In the last session, we started reasoning about connections between factors, and generated a link circle displayed on the screen. In this session, we’ll use the understanding we’ve built through this exercise. We will go from a link circle to a graph structure, to better understand the structure and behavior of the system, and look at multiple links in a row. We’ll start with the basic model we’ve discussed at the beginning.*

*We’ll start with the basic model we’ve discussed at the beginning.*

*[The facilitator tapes up a large sheet of paper containing the simple graph, and projects a slide with a list of the final factors.]*

*Here's how we're going to add a link.*

*[Choose* a factor that is on the final list of factors but not yet added to the simple graph. *]*

*We'll add factors from the final list we've identified. The links in the integrated link circle are a suggestion, but additional links that were not identified in the link circle can be added to the graph, and all links from the link circle need not be added to the graph.*

*Each group will generate their own graph, and we will combine the insights into a single graph in the next session. Please discuss connections you want to add with members of your group - if you disagree, and you can't find an easy resolution, please write it down on the side and share it with the group later.*

*Remember that in addition to factors that have a direct relationship with the main factor (kidney health), there may also be factors that are connected to the main factor via intermediate factors. For example,*

*[show an example]*

*You will have 15 minutes to work on this exercise in your small groups. We'll be walking around to offer assistance, let us know if you have any questions/*

*[2 minute warning]*

*You have 2 minutes left. In 2 minutes, we'll ask one person from each group to describe components of the graphs you've generated.*

**References:**

1. This script is modified from: *Scriptapedia: Model Review*[12] and *Scriptapedia: Creating Causal Loop Diagram from Connection Circles*[13] and *Scriptapedia: Causal Mapping in Small Groups*[14]

#### Unified graph creation script

**Time:** 1:30–2:00 pm
**Duration:** 30 min
**Format:** Plenary

**Materials:**
- Projector

**Roles:**

- Facilitator: introduces and facilitates this script
- Modeler: creates the integrated graph in VenSim
- Process coach: supports small groups groups who may require assistance

**Inputs:**
- One graph generated per small group

**Outputs:**
- One integrated graph

**Vocabulary: [terminology used in this script: other GMB terminology]**
- Factor: variable
- Link: causal link
- Link type: link polarity
- Positive link: positive link polarity
- Negative link: negative link polarity
- Graph: causal loop diagram
- Simple graph: concept model

**Steps:**
1. The modeler projects the simple graph in VenSim using the projector, and each small group keeps their graph at their table. The facilitator informs the group that we will now be integrating the graphs that each group has created into a shared graph.
2. The facilitator has the small groups share a component of their graph that they would like to add to the integrated graph, in a round-robin fashion. An addition may involve one or more factors that are not yet added to the integrated graph along with links, or just adding links.
    a. As each addition is shared, the facilitator prompts the participant sharing to provide a brief description or rationale for the addition.
    b. As each addition is suggested, the modeler includes this in the graph on VenSim that is projected onto the screen in order to visualize what is meant. The facilitator checks with other group members if they agree with the proposed addition.
    c. If someone disagrees, the facilitator can ask for clarification and try to determine what the group thinks the relationship should be. Remind the group that if a discussion goes on too long, the group can choose to temporarily 'park' this item and continue with another part of the graph.
    d. As the groups continue adding from the graphs that they have created in the small groups, the facilitator suggests that they may also begin to suggest additions that were not present in the small groups' graphs.
3. The facilitator ends the session by noting that the graph represents a combination of each small group's graph and understanding of the connections between the factors.
4. The recorder captures the products of the process photographically.

**References:**

1. This script is modified from: *Scriptapedia: Causal Mapping in Large Group*[15] (and from our link circle integration script – to check whether any of the references for that are also used here)
   [Note added: While the Causal Mapping in Large Group script from Scriptapedia was the only script that was directly referenced in creating this script, we did additionally refer to our Link Circle Integration Script.]

***Facilitator 1:***

*We will now be integrating graphs that each group has created into a shared graph. We'll ask groups to share additions to the graph one component at a time. A component is a part of the graph that captures a meaningful mechanism - it can be a single factor, but it can also be a loop or a path of multiple factors, the way the basic graph is. As you're reading it out, we'll ask you to give a brief description of the mechanism and why you want to add it.*

*We'll add them in VenSim and display them on the screen.*

*[As factors are added]*

- *Does everyone agree with this addition?*
- *Why don't you agree? What would you change?*
- *What does everyone else think?*
- *It seems this is a more challenging question - let's come back to it later, and continue with other parts of the graph.*
- *You may also begin to suggest additions that were not present in the small groups' graphs.*

*[end of session] The graph represents a combination of each small group's graph and understanding of the connections between the factors.*

*We'll now take a short break. When we come back, we're going to analyze the graph we created.*

#### Break

**Time:** 2:00–2:15 pm
**Duration:** 15 min

**Materials:**

- Printer
- A4 sheets of paper

**Roles:**

- Additional team member: Prints out the graph created in the previous script. This document is printed out on A4 sheets of paper, with each graph spread out across 4 sheets of paper. The sets of 4 sheets of paper are taped together to make graph printouts. Six copies of this are prepared.

#### Loop and path identification script

**Time:** 2:15–2:45 pm
**Duration:** 30 min
**Format:** Small group

**Materials:**
- Printed out graphs (one per small group) – 4 A4 sheets of paper taped together for each graph
- Markers
- Projector

**Roles:**
- Facilitator: introduces the activity, including providing an example path and loop
- Process coach: supports small groups groups who may require assistance

**Inputs:**
- Graph generated by the group

**Outputs:**
- 3 most important loops/paths identified per small group

**Vocabulary: [terminology used in this script: other GMB terminology]**
- Factor: variable
- Link: causal link
- Link type: link polarity
- Positive link: positive link polarity
- Negative link: negative link polarity
- Graph: causal loop diagram
- Loop: feedback loop
- Simple graph: concept model

**Steps:**
1. The facilitator introduces that in this activity, the group will be exploring and improving the graph to better understand how the system currently works. Particularly, the group will be exploring the presence of meaningful paths and loops in the graph. The facilitator defines paths and loops, and provides an example from the simple graph for each. The examples are visually shown through displaying the graph via the projector and pointing out the examples.

a. The facilitator defines that a path is when we can identify a starting factor and an ending factor, and there is a pathway of links that we can follow to influence the ending factor if we start at the starting factor. Note that in a path, it is important to have a continuous pathway of links, but the links can be a mix of positive and negative links. For instance, in the simple graph, there is a path from 'Motivation to improve health' to 'Receiving timely care'.
b. The facilitator defines that a loop is when there is a path from a factor back to itself; that is, where the starting factor and the ending factor are the same. For instance, in the simple graph, there is a loop from 'Kidney health' back to 'Kidney health', with intermediate factors of 'Motivation to improve health', 'Seeking care', and 'Receiving timely care'.

2. The facilitator asks whether the group has any questions about the examples shared, and reiterates that the group will be exploring paths and loops within the graph they have created. The facilitator asks the participants to return to their small groups, and each small group is handed a printed out graph and markers. The facilitator asks each small group to identify meaningful paths and loops, and to identify the three most meaningful paths/loops to share with the larger group.
    a. The facilitator mentions that we would like to focus on paths/loops that contain 3-5 factors, but the groups can identify paths/loops of more factors if they seem important.
    b. The facilitator adds that while identifying paths or loops, participants may find that they need to add/remove links or otherwise modify the graph. The facilitator mentions that the small groups should keep track of these proposed modifications as well, and we'll discuss them as a larger group when we come back together.
3. The small groups are given 15 minutes to identify paths/loops and to choose the three most important ones that they would like to share with the larger group. When 3 minutes remain, the groups are given a warning of the time remaining, and are asked to focus on deciding which loops/paths they would like to share out.

***Facilitator 2:***
*During this session, we will be exploring and improving the graph to better understand how the system currently works. We will focus on the presence of meaningful paths and loops in the graph.*

*[Show a path and a loop] This is an example of a path [draw, read out]. This is an example of a loop [draw, read out].*

1. *<u>A path</u> is when we can identify a starting factor and an ending factor, and there is a pathway of links that we can follow to impact the ending factor if we start at the starting factor. Note that in a path, we can have both positive and negative links. For instance, in the simple graph, there is a path from a steady job to good kidney health.*
2. *<u>A loop</u> is when there is a path from a factor back to itself; that is, where the starting factor and the ending factor are the same. For instance, in the simple graph, there is a*

*loop from good kidney health back to good kidney health, with intermediate factors of steady job, good health care, and healthy nutrition.*

*Are there any questions about the examples shared?*

*Now, we'll ask that you return to your small groups. Each group will be given a copy of the graph, and we'll ask you to identify loops and paths and share 3 most important ones to a group. Think about the meaning of those paths - why do you think they are important? Are any loops and paths you find surprising?*

*You'll have 15 minutes.*

*While identifying paths or loops, you may find that they need to add/remove links or otherwise modify a graph. Please keep track of these proposed modifications as well, and we'll discuss them as a larger group when we come back together.*

*[3 minutes remaining]. There are 3 minutes remaining. As you're starting to wrap up, think about the most important paths/loops you'd like to share out.*

**References:**

1. This script is modified from: *Scriptapedia: Creating Causal Loop Diagram from Connection Circles*[13]

#### Loop and path integration script

**Time:** 2:45–3:15 pm
**Duration:** 30 min
**Format:** Plenary

**Materials:**

- Projector
- iPad

**Roles:**

- Facilitator: Facilitates the sharing of the loops/paths and modifications, annotates the loops/paths and adds the modifications via iPad screen projected
- Modeler: Adds the modifications suggested to the graph in VenSim
- Recorder: Captures the loops, paths, and modifications suggested by the small groups

**Inputs:**

- From each small group – 3 most meaningful paths or loops
- From each small group – proposed changes to the graph

**Outputs:**

- Modified graph annotated with meaningful paths/loops

**Vocabulary: [terminology used in this script: other GMB terminology]**

- Factor: variable
- Link: causal link
- Link type: link polarity
- Positive link: positive link polarity
- Negative link: negative link polarity
- Graph: causal loop diagram
- Loop: feedback loop

**Steps:**

1. The facilitator expresses that the small groups will be sharing out the loops and paths that they identified, but first we'll explore any proposed modifications to the graph that came up in their discussions.
2. The modeler projects the graph on VenSim. The facilitator asks the groups if they each have any modifications (additions or deletions of links) to suggest. As the modifications are shared, the modeler demonstrates the modification on VenSim so it is visual to the entire group, and the facilitator asks whether the entire group agrees on the change.
3. Once all modifications have been shared, the iPad screen is shared using the projector, with an image of the modified graph that can now be annotated with loops/paths.
4. The facilitator guides the small groups to share out the loops/paths they identified, in a round robin format, where each group shares one loop/path at a time. As they share the loop/path. The facilitator prompts each small group to share a brief interpretation of the loop/path, in addition to naming the factors that form the loop/path.
5. As a participant shares a loop/path/modification, the modeler annotates (via iPad) the graph that is projected, so that the entire group can see the loop/path that is being named. Each new loop or path is annotated in a different color. For every suggestion, the facilitator asks the entire group whether the loop/path makes sense to everyone.
6. The facilitator summarises by mentioning the insight into the graph that has been obtained through this exploration of loops, paths, and modifications.

***Facilitator 1:***

*The small groups will now be sharing out the loops and paths that they identified, as well as any proposed modifications to the graphs that came up in their discussions.*

*As you share, I'll ask you to briefly describe the connections.*

*[Share]*

*Now, after exploration of loops, paths, and modifications to the graph, we have a deeper insight into the graph.*

**References:**

1. This script is modified from: [No specific script was used here]
   [Note added: While no script from Scriptapedia was directly referenced in creating this script, we did refer to the previous scripts that we created.]

##### Coffee break

**Time:** 3:15–3:30 pm
**Duration:** 15 min

**Materials:**
- Printer + A4 sheets of paper
- Tape

**Roles:**
- Additional team member: Prints out the annotated (with loops/paths), modified graph on sheets of paper, such that a single graph is across 4 A4 sheets of paper. Tapes the 4 sheets of paper together, and creates a total of 6 copies of the graph.
- Facilitator / Modeler: Begin to identify a few key insights and themes that have arisen in the workshop so far, to incorporate into the final wrap-up script. Identify a place in the graph to add a dot, as an example for the dot exercise.

#### Session 4

##### Dot exercise for interventions script

**Time:** 3:30–4:00 pm
**Duration:** 30 min
**Format:** Plenary

**Materials:**
- Projector
- Adhesive dots
- Printed out graphs

**Roles:**
- Facilitator: Introduces dot exercise and facilitates reflection
- Modeler: Adds the dots added by participants to a figure of the graph on Google Slides
- Recorder: Records the locations + reasons for dot placement

**Inputs:**
- Graph with modifications and annotated with meaningful loops/paths

**Outputs:**
- Graph with dots added, one dot per participant

**Vocabulary: [terminology used in this script: other GMB terminology]**

- Factor: variable
- Link: causal link
- Graph: causal loop diagram

**Steps:**

1. The printed out graphs (annotated with loops/paths) are evenly distributed in front of the participants, so that each participant is close to and is able to see one of the printouts. The graph is also projected onto the screen. The facilitator states that since the group has developed a deeper understanding of the system through building the graph, they will now select areas of the graph where changes could be made to help patients.
2. The facilitator gives every participant 1 adhesive dot, and tells participants that they will each get to place their dot on one of the printed out graphs in front of them, beside the area in the graph where they most think that a change could be helpful to patients, through eventually impacting the main factor of kidney health.
    a. A change could either be to a factor or a link.
    b. A factor could be changed directly by increasing or decreasing it. However, it may be important to think about factors that impact the factor we are changing, such as upstream factor.
    c. A link could be changed by strengthening or weakening it.
3. The facilitator provides an example of where they could place a dot, as identified in the previous coffee break.
4. The facilitator states that they will give the participants 5 minutes to think about where they want to place their dot, and that when they are ready, they can place their dot on one of the graphs in front of them. When the group returns after 5 minutes, each person will get the chance to share where they placed their dot and why.
5. The facilitator indicates the beginning of the 5 minutes, and gives the group a 1 minute warning. During the 1 minute warning, the facilitator reminds participants that they will get the chance to share with the larger group where they placed their dot and why.
6. When the 5 minutes are over, the facilitator asks the room to go in a circle and for each person to share where they placed their dot and why. The modeler adds each person's dot, as they share, to the graph on the screen. Each person is given ~1 minute to share.
7. The facilitator summarizes any patterns that have emerged in where dots have been added to the graph, and therefore where participants would most like to see interventions.

**References:**

1. This script is modified from: *Scriptapedia: Action Ideas*[16] and *Scriptapedia: Dots*[17]

***Facilitator 3:***

*Now that we have developed a deeper understanding of the system through building the graph, we'll now ask you to select areas of the graph where changes could be made to help patients.*

*Those changes can be coming from the health system, government, organizations, etc - you're welcome to point out changes you'd like to see, say, from Stanford, but you don't have to.*

*We'll give each of you 1 sticker, and ask you to place it on a graph. For example, if I am someone who believes that [UPDATE: transportation] is perhaps the most important barrier, I can place my dot near this factor, or near one of the links to indicate what process exactly I'd like to see changed, such as the way in which it impacts [UPDATE: getting to appointments].*

*In some cases, it may be important to think about factors that impact the factor we are changing, such as upstream factor.*

*We will now give you 5 minutes to individually think about where you'd like to place the sticker. When you're ready, you can place it on the graph in front of you. We will then go around and give everyone a chance to describe what they chose and why.*

*[5 minutes later]*

*We're now going to go around and give everyone a chance to describe what they chose and why. We will have about a minute per person, and we will have a chance later to share additional thoughts if you have more to say. Would you like to get us started?*

*[Each person is given ~1 minute to share]*

*[Summarizing patterns]*
- *Ok, so it sounds like many of you agree that xxx is very important, and we've heard that xxx.*
- *It seems that the part of the graph related to xxx received a lot of votes, and the part related to xxx less so*
- *…*
- *It appears that many of you agree that xxx [area of graph] should be a focus area for interventions*

#### Reflection and wrap-up script

**Time:** 4:00–4:30 pm
**Duration:** 30 min
**Format:** Plenary

**Materials:**
- Reflection cards
- Pens
- Projector

**Roles:**
- Facilitator: Wraps up the session and thanks the participants

- Modeler: Projects the final graph (with dots added)

**Inputs:**
- NA

**Outputs:**
- Reflections (shared verbally and/or through reflection cards)

**Steps:**
1. The modeler projects the final graph with dots added, indicating the final causal graph created by the group with a visual representation of places in the graph where participants would most like to see interventions.
2. The facilitator calls attention to the final graph created, indicating that it is a combination of the knowledge and experiences that each participant has brought to the workshop. The facilitator mentions a few key themes and insights that have arisen from the creation of the graph.
3. The modeler hands each participant a reflection card and pen. The facilitator states that 5 mins will be provided for each participant to reflect on the experiences of the day, and they may write down any reflections on the reflection card provided to them. At the end of the 5 mins, the participants may share their reflections verbally, or may share their reflection card with the research team.
4. After 5 minutes, the facilitator asks the participants if anyone would like to share any of their reflections, and asks 4-5 people to share their reflections. The participants are also given the option to hand their reflection card to the facilitator.
5. The facilitator wraps up the workshop by thanking the participants and mentioning any next/future steps.

**References:**
1. This script is modified from: [NA for now]
   [Note added: While no script from Scriptapedia was directly referenced in creating this script, we did refer to the previous scripts that we created.]

***Facilitator 1:***
*We've come to the end of our workshop - we'd like to thank all of you for participating, it was a long day, and we learned a lot from all of you.*

*Here's the final product from today [show displayed graph]. It is a combination of the knowledge and experiences that each of you has brought to the workshop.*
- *[describe insights from the graph]*
- *The graph we've generated shows that many factors are important in taking care of kidney health, [list themes]*
- *We've captured how xxx is important in maintaining kidney health through xxx and xxx, and how xxx plays a role in that*

*In the last 20 minutes of the workshop, we'd like to give you a chance to reflect on this experience and share any reflections you'd like with the group. Each of you got a reflection card - for the next 5 minutes, think about important takeaways from today, things that have been important to you or that you'd like to share with others. You can share the card with the team, or if you'd like to keep your reflection to yourself, you can hold onto your card.*

*If anyone would like to share it with the group, we will have about [10?] minutes.*

*[After sharing]*

*Thank you everyone, this ends our workshop. Again, we would like to thank all of you for spending your Saturday with us - your knowledge and insights are incredibly important, and will be shared with Stanford Health Care and with researchers working to improve care for chronic kidney disease patients.*

*The study team will stay here for [xxx] more minutes if you have any questions or any additional thoughts you'd like to share.*

#### End (with 30 min buffer)

### Supplement 6: Causal Loop Diagram Refinement Steps

During post-workshop refinement of the CLD, the modeling team iterated on the CLD generated from the workshop. The changes made at this stage are listed in Supplemental Table S4.

Cascading changes have not been included in Supplemental Table S4, which are changes that follow immediately from any of the changes in this log. For instance, if removal of a variable results in removal of a link to or from that variable, then removal of the link is considered a cascading change and is not included. Additionally, when it is mentioned that a connection was discussed by participants, this could be either at the GMB workshop, during focus groups, or both.

Supplemental Table S4: Causal Loop Diagram Changes

| Change Number | Change Type | Original Element | Modified Element | Change Description and Rationale |
|---|---|---|---|---|
| 1 | Variable added | – | *Having a job* | This variable was added based on participant discussions regarding the impact of employment on having insurance (and therefore ability to access care), and on having time for CKD care. Although it wasn't initially chosen to include in the graph during variable selection, the modeling team decided to include it in the modified version due to subsequent participant discussions. |
| 2 | Variable added | – | *Having time* | This variable was added since the amount of time required to manage care was mentioned by participants. Additionally, to better capture how community support impacts engagement with clinical care, through having time as a mediating variable. Having time as the mechanism for how community support impacts care management was discussed by participants but not formalized, since a variable representing having time was not selected during variable selection in the GMB workshop. Thus, while modifying the graph, the modeling team |

| | | | | |
|---|---|---|---|---|
| | | | | decided to introduce this variable. |
| 3 | Variable removed | *Continuity of care* | – | This variable was removed since it has semantic overlap with *Coordination of care* and *Receiving timely care* in the participant-created graph, and is only linked to those two variables. |
| 4 | Variable removed | *Caretaking duties* | – | Since caretaking duties is sparsely linked to the remainder of the graph, only through *Community support* and *Seeking care* (which is converted to *Engagement with clinical care*, and since participant discussions represented *Caretaking duties* primarily in the context of how much time can be taken by such duties, and how much community support can lessen the time burden of caretaking duties, we remove this variable and incorporate the knowledge formerly represented by this variable through the link added between *Community support* and *Having time*. This is indicated in another change in this log, where that link is added. |
| 5 | Variable renamed | *Receiving timely care* | *Receiving timely CKD care* | This variable was renamed to specify that receiving timely care is specifically in the context of timely CKD care, to distinguish it from holistic, preventive, and non-CKD care. |
| 6 | Variable renamed | *Co-occurring chronic illnesses* | *Non-CKD disease burden* | This variable was renamed to expand its scope to capture not just non-CKD chronic diseases, but rather, to capture complete health status except CKD. |
| 7 | Variable renamed | *Knowledge about CKD management* | *Knowledge about CKD and management* | This variable was renamed to clarify that knowledge may be about CKD itself (disease process, etc.) or about strategies to manage CKD. |

| | | | | |
|---|---|---|---|---|
| 8 | Variable renamed | *Access to nutrition support* | *Access to nutritionist* | This variable was renamed to clarify that it represents access to a nutritionist specifically, rather than the more expansive definition that could capture nutrition-related support from community support. Following this change and to better represent the participant discussions that community support can improve nutritional support more broadly rather than relating to a nutritionist, the next two changes remove the link from *Community support* to *Access to nutritionist* and add a link from *Community support* to *Eating healthy and getting exercise*. |
| 9 | Variable renamed | *Doctor trust* | *Doctor trustworthiness* | We changed the name of this variable to more closely represent the semantic meaning of this variable in participant discussions. The focus of this variable is on the doctor's behaviors and actions which signal trustworthiness to patients. |
| 10 | Variables merged | *Access to health care services*, *Receiving timely CKD care* | *Receiving timely CKD care* | Due to the semantic overlap between *Access to health care services* and *Receiving timely CKD care*, we assumed that *Access to health care services* is fully captured by *Receiving timely CKD care*. |
| 11 | Variables merged | *Getting exercise*, *Eating healthy* | *Eating healthy and getting exercise* | We noted that *Eating healthy* and *Getting exercise* had many of the same links to and from them. Additionally, the links that they did not share made semantic sense to add and had been discussed by participants. Thus, the modeling team chose to combine these two variables into a single variable to capture the system overlap—shared causal links and shared role in the graph—between them. |
| 12 | Variables merged | *Coordination of care*, *Preventative* | *Coordinated and holistic care* | We combined *Coordination of care* and *Preventive and holistic care* due to the overlap in links to and from these |

| | | | | |
|---|---|---|---|---|
| | | *and holistic care* | | variables in the participant-created graph. Additionally, the links that didn't have overlap were plausible to add and had often been discussed by participants. |
| 13 | Variables merged | *Seeking care*, *Treatment compliance* | *Engagement with clinical care* | We combined *Seeking care* and *Treatment compliance* into the semantically broader variable *Engagement with clinical care* to express both seeking care and continuing to remain engaged with care. |
| 14 | Link removed | *Preventative and holistic care → Kidney health* | – | The variable *Preventive and holistic care* was modified into *Coordinated and holistic care*. Yet, we chose to remove the link from *Coordinated and holistic care* to *Kidney health* since we determined that *Coordinated and holistic care* impacts *Kidney health* indirectly via mediators rather than directly. Mediators in this case could include *Receiving timely CKD care*, *Non-CKD disease burden*, or *Access to nutrition support* and *Eating healthy and getting exercise*. |
| 15 | Link removed | *Co-occurring chronic illnesses → Coordination of care* | – | Both variables in this link had their names changed. However, we chose to remove the link between the changed name variables (from *Non-CKD disease burden* to *Coordinated and holistic care*), to more closely represent participant discussions. The link was initially added during the GMB workshop since participants reasoned that patients with more chronic illnesses may be in more contact with medical care systems. However, participants also discussed that having a greater disease burden (CKD and otherwise) and therefore more contact with medical care systems does not imply a greater coordination of care. Thus, we removed this link to represent |

| | | | | |
|---|---|---|---|---|
| | | | | this discussion. |
| 16 | Link added | – | *Having time → Engagement with clinical care* | Related to the change of adding the variable *Having time*, a link from *Having time* to *Engagement with clinical care* was added. This connection was discussed by participants, yet was not added in the participant-generated graph since *Having time* was not selected as a variable during the variable selection phase of GMB. |
| 17 | Link removed | *Community support → Access to nutrition support* | – | Since we renamed the variable *Access to nutrition support* to *Access to nutritionist* to reflect the specific access to a nutritionist within the broader access to nutrition support (which could include community-based nutrition support), we removed this link. Participants discussed how community support can impact the ability to eat healthy, rather than the ability to access a nutritionist. Thus, removing this link and adding the link from *Community support* to *Eating healthy and getting exercise* (as added in another change) more closely represents participant discussions. |
| 18 | Link removed | *Access to health care services → Seeking care* | – | This link was initially added to reflect patient hesitancy to seek care or go to a doctor if they do not have access to care. However, since we combined the variable *Access to health care services* with the variable *Receiving timely CKD care*, the link from *Receiving timely CKD care* to *Engagement with clinical care* (which is the variable name that *Seeking care* was renamed to) no longer captures this connection discussed by participants—so, we removed this link. Patient hesitancy to engage with or seek care if they do not have access to care, as discussed by participants, is instead captured via the |

| | | | | |
|---|---|---|---|---|
| | | | | link added from *Insurance coverage* to *Engagement with clinical care* (added in another change). |
| 19 | Link polarity changed | *Eating healthy* → (+) *Co-occurring chronic illnesses* | *Eating healthy and getting exercise* → (-) *Non-CKD disease burden* | We believe that the polarity for this link was incorrectly marked during the GMB workshop. Per participant discussions and our reasoning, an increase in *Eating healthy and getting exercise* would cause a decrease in *Non-CKD disease burden*, and vice-versa. |
| 20 | Link removed | *Treatment compliance* → *Kidney health* | – | The variable *Treatment compliance* was converted to *Engagement with clinical care*, yet we removed the link from *Engagement with clinical care* to *Kidney health*. We believe that this direct link in the participant-generated graph is best captured by indirect links (via *Receiving timely CKD care*, for instance). |
| 21 | Link added | – | *Insurance coverage* → *Engagement with clinical care* | We added a link from *Insurance coverage* to *Engagement with clinical care*, since although links from *Insurance coverage* to *Seeking care* or *Treatment compliance* were not added to the participant-generated graph, participants discussed that a lack of insurance or access to care would dissuade them from seeking or engaging with care. This link was added to represent these participant discussions. |
| 22 | Link added | – | *Insurance coverage* → *Receiving timely CKD care* | The connection between insurance and receiving care was discussed by participants. |
| 23 | Link added | – | *Knowledge about CKD and management* → *Motivation to improve* | Although this link was not added to the participant-generated graph, participants discussed the connection between these factors. |

| | | | | |
|---|---|---|---|---|
| | | | *health* | |
| 24 | Link added | – | *Having a job → Money or income* | *Having a job* is a new variable that was not present in the participant-generated graph. This link was added based on participant discussions. |
| 25 | Link added | – | *Community support → Having time* | *Having time* is a new variable that was not present in the participant-generated graph, so links to and from it were not present. This link was added to the modified graph based on participant discussions. |
| 26 | Link added | – | *Having time → Eating healthy and getting exercise* | *Having time* is a new variable that was not present in the participant-generated graph. Participants discussed the importance of having time for engaging in healthy behaviors such as eating healthy and getting exercise. |
| 27 | Link added | – | *Coordinated and holistic care → Access to nutrition support* | Although this link was not added to the participant-generated graph, participants discussed the importance of a holistic approach for gaining access to nutrition support. |
| 28 | Link added | – | *Having a job → Having time* | Both the variables in this link were not present in the participant-generated graph. However, participants discussed the causal connections between employment, having time, and managing their care. Specifically, they discussed that having a job can be an impediment to managing CKD care, due to care being time-consuming. Thus, this link was added to reflect this discussion. |
| 29 | Link added | – | *Engagement with clinical care → Knowledge about CKD and management* | Participants discussed how engaging with their care can impact their knowledge about CKD and management, so we added this link to the graph. We believe this link was less clear previously, when *Engagement with clinical care* was instead represented by the variables *Seeking* |

| | | | | |
|---|---|---|---|---|
| | | | | *care* and *Treatment compliance*. |
| 30 | Link added | – | *Having a job → Insurance coverage* | Since *Having a job* is a new variable added, this link was not previously present in the participant-generated graph. However, participants discussed the connection between employment and insurance coverage. |
| 31 | Link added | – | *Engagement with clinical care → Having time* | Participants discussed that engaging with care is time-consuming, indicating this link. |
| 32 | Link added | – | *Eating healthy and getting exercise → Having time* | Participants discussed that eating healthy and getting exercise is time-consuming, indicating this link. |
| 33 | Link added | – | *Community support → Eating healthy and getting exercise* | Participants discussed how community support can impact the ability to eat healthy, such as through providing support with CKD-friendly dining options. This link addition is related to two other changes made, where the link from *Community support* to *Access to nutrition support* was removed as *Access to nutrition support* was renamed to *Access to nutritionist*. This link added thus retains the participant-discussed relationship between community support and ability to eat healthy. |
| 34 | Link added | – | *Knowledge about CKD and management → Community support* | This link was added to represent participant discussions and experiences about how knowledge about CKD and management can lead to an increase in availability or access of community support. For instance, through knowing about support groups. |
| 35 | Link added | – | *Community support → Knowledge about CKD and* | We added this link to represent participant discussions of how community support can lead to an increase in knowledge about CKD and management, through sharing |

| | | | | |
|---|---|---|---|---|
| | | | *management* | knowledge, such as through support groups and the types of discussions that occurred during our focus groups and workshops, where participants were able to share knowledge with each other. |
| 36 | Link added | – | *Doctor trustworthiness → Knowledge about CKD and management* | This link was added to represent that doctor trustworthiness can increase the knowledge that a patient has about CKD and management through the doctor directly providing relevant information to the patient. |

2. Hovmand P, Rouwette E, Andersen D, et al. Scriptapedia: a handbook of scripts for developing structured group model building sessions.

3. Social Determinants of Health. *Public Health Professionals Gateway*, https://www.cdc.gov/public-health-gateway/php/about/social-determinants-of-health.html.

4. Andersen D, Richardson G. Scriptapedia/Presenting the Reference Mode. *Wikibooks, open books for an open world*, https://en.wikibooks.org/wiki/Scriptapedia/Presenting_the_Reference_Mode.

5. Richardson GP. Scriptapedia/Concept Model. *Wikibooks, open books for an open world*, https://en.wikibooks.org/wiki/Scriptapedia/Concept_Model.

6. Team NPE. Social Determinants of Health and Chronic Kidney Disease. *National Kidney Foundation*, https://www.kidney.org/kidney-topics/social-determinants-health-and-chronic-kidney-disease.

7. Andersen DF, Richardson GP. Scriptapedia/Variable Elicitation. *Wikibooks, open books for an open world*, https://en.wikibooks.org/wiki/Scriptapedia/Variable_Elicitation.

8. Andersen DF, Richardson GP. Scriptapedia/Connection Circle. *Wikibooks, open books for an open world*, https://en.wikibooks.org/wiki/Scriptapedia/Connection_Circle.

9. *Vensim PLE*. Ventana Systems, https://vensim.com/.

10. Molloy J. Learning about connection circles. *The Systems Thinker*, https://thesystemsthinker.com/learning-about-connection-circles/.

11. Ellson J, Gansner ER, Koutsofios E, et al. Graphviz and dynagraph — static and dynamic graph drawing tools. *Mathematics and Visualization* 2004; 127–148.

12. Scriptapedia/Model Review. *Wikibooks, open books for an open world*, https://en.wikibooks.org/wiki/Scriptapedia/Model_Review.

13. Hovmand P, Kraus A. Scriptapedia/Creating Causal Loop Diagram from Connection Circles. *Wikibooks, open books for an open world*, https://en.wikibooks.org/wiki/Scriptapedia/Creating_Causal_Loop_Diagram_from_Connection_Circles.

14. Ballard E. Scriptapedia/Causal Mapping in Small Groups. *Wikibooks, open books for an open world*, https://en.wikibooks.org/wiki/Scriptapedia/Causal_Mapping_in_Small_Groups.

15. Ballard E. Scriptapedia/Causal Mapping in Large Group. *Wikibooks, open books for an open world*, https://en.wikibooks.org/wiki/Scriptapedia/Causal_Mapping_in_Large_Group.

16. Scriptapedia/Action Ideas. *Wikibooks, open books for an open world*, https://en.wikibooks.org/wiki/Scriptapedia/Action_Ideas.

17. Scriptapedia/Dots. *Wikibooks, open books for an open world*, https://en.wikibooks.org/wiki/Scriptapedia/Dots.
